# Development of Childhood Asthma Prediction Models using Machine Learning Approaches

**DOI:** 10.1101/2021.03.31.21254678

**Authors:** Dilini M. Kothalawala, Clare S. Murray, Angela Simpson, Adnan Custovic, William J. Tapper, S. Hasan Arshad, John W. Holloway, Faisal I. Rezwan, on behalf of STELAR/UNICORN investigators

**Author notes:** Co-authors contributed equally (John W. Holloway, Faisal I. Rezwan). **Correspondence to:** Name: Professor John W Holloway, Address: Duthie Building, MP808, Southampton General Hospital, SO16 6YD, United Kingdom. **Funding:** This work was supported by the National Institute for Health Research through the NIHR Southampton Biomedical Research Centre and a University of Southampton Presidential Research Studentship. Replication analysis in MAAS was supported by the Medical Research Council as part of UNICORN (Unified Cohorts Research Network): Disaggregating asthma MR/S025340/1. Angela Simpson and Clare Murray are supported by the NIHR Manchester Biomedical Research Centre. **Conflicts of interest:** The authors have no funding relationships or conflicts of interest related to this article to disclose. **Data availability:** Access to data from the Isle of Wight Birth Cohort can be made available upon request. Further information can be found at www.allergyresearch.org.uk/ or from: Cohort Profile: The Isle Of Wight Whole Population Birth Cohort (IOWBC). Int J Epidemiol. 2018 Aug 1;47(4):1043-1044i. doi: 10.1093/ije/dyy023. Source code for the development and use of the prediction models can also be made available upon request.

## Abstract

**Background:** Wheeze is common in early life and often transient. It is difficult to identify which children will experience persistent symptoms and subsequently develop asthma. Machine learning approaches have the potential for better predictive performance and generalisability over existing childhood asthma prediction models.

**Objective:** To apply machine learning approaches for predicting school-age asthma (age 10) in early life (Childhood Asthma Prediction in Early life, CAPE model) and at preschool age (Childhood Asthma Prediction at Preschool age, CAPP model).

**Methods:** Data on clinical symptoms and environmental exposures were collected from children enrolled in the Isle of Wight Birth Cohort (N=1368, ∼15% asthma prevalence). Recursive Feature Elimination (RFE) identified the optimal subset of features predictive of school-age asthma for each model. Seven state-of-the-art machine learning classification algorithms were used to develop the models and the results were compared. To optimize the models, training was performed by applying 5-fold cross-validation, imputation and resampling. Predictive performances were evaluated on the test set and externally validated in the Manchester Asthma and Allergy Study (MAAS) cohort.

**Results:** RFE identified eight and 12 predictors for the CAPE and CAPP models, respectively. The best predictive performance was demonstrated by a Support Vector Machine (SVM) algorithm for both the CAPE model (area under the receiver operating curve, AUC=0.71) and CAPP model (AUC=0.82). Both models demonstrated good generalisability in MAAS (CAPE 8YR=0.71, 11YR=0.71, CAPP 8YR=0.83, 11YR=0.79).

**Conclusion:** Using machine learning approaches improved upon the predictive performance of existing regression-based models, with good generalisability and ability to rule in asthma.

## INTRODUCTION

Childhood asthma is highly heterogeneous, with numerous factors contributing towards its development, persistence and severity^1-3^. Despite approximately 80% of asthmatic children developing symptoms (such as wheeze) before the age of six, these clinical symptoms are neither universally present in early life among all future asthmatics nor specific to asthma^4^. With the added difficulty of making an objective asthma diagnosis before the age of five, both under-treatment and over-treatment of wheezing disorders are common in early life^5,6^.

The ability to predict the development of school-age asthma can help to identify high-risk pre-school children and distinguish them from children whose symptoms are likely to be transient. Furthermore, early prediction of asthma susceptibility will be critical for the successful implementation of potential primary prevention strategies to reduce the risk of developing asthma.

A recent systematic review identified twenty-one regression-based models for predicting childhood asthma^7^. However, none of these models have been implemented into standard clinical practice; this is possibly due to their relatively poor predictive power, poor generalisability upon independent validation and need for specialised clinical testing. The review further proposed that regression-based methods for predicting childhood asthma may have been exhausted, with the identified models offering similar predictive power to each other and being unable to be significantly improved upon^7^.

Machine learning approaches have increasingly been applied to address a wide range of healthcare problems due to their ability to integrate large quantities of heterogeneous data, handle complex interactions between variables and identify patterns within data compared to traditional statistical methods^8^. Particularly for disease prediction, where interactions between biological variables are complex, machine learning approaches have the potential to identify novel predictors which were previously overlooked by regression-based approaches^8-10^. Furthermore, the lack of application of methods to reduce model overfitting may address the poor generalisability of existing prediction models in independent populations. Machine learning approaches have shown promise in predicting a variety of clinical asthma outcomes, phenotypes and decisions^11-15^. However, there have been few reports of machine learning methods being applied for either the diagnostic or prognostic prediction of childhood asthma development^16-20^.

This study aimed to explore whether machine learning approaches can offer an improvement over traditional regression-based methods for predicting school-age asthma. To predict school-age asthma at 10 years, two models, the Childhood Asthma Prediction in Early-life (CAPE) and Childhood Asthma Prediction at Preschool-age (CAPP) models were developed within a general population based cohort using information available from the first two years and first four years of life, respectively.

## METHODS

### Developmental Study Population

Data was obtained from 1456 individuals enrolled in the Isle of Wight Birth Cohort (IOWBC). Study recruitment and participant details have been previously described^21^ (see Supplementary Methods).

#### Prediction outcome

School-age asthma, evaluated at age 10, was defined as “a doctor diagnosis of asthma ever and at least one episode of wheezing or use of asthma medication in the last 12 months”. Only individuals with a reported asthma status at the 10-year follow-up were included in the analyses.

#### Candidate predictors

Reported risk factors associated with childhood asthma in the literature were used to identify 54 candidate predictors for which data was available in the IOWBC (Table E1) and underwent pre-processing (see Supplementary Methods). Candidate predictors included data on subject demographics, lifestyle, clinical symptoms of allergy and asthma and environmental exposures collected across three time points: at birth (prenatal and perinatal data), early life (combined exposure at either the 1-year or the 2-year follow-ups) and at preschool age (4-year follow-up).

### Model development

All stages of model development were performed independently for the CAPE and CAPP models.

#### Feature selection

For each model, feature selection was performed on the complete dataset for all available candidate predictors (without any missing values) using Recursive Feature Elimination (RFE) with a random forest algorithm, using 5-fold cross-validation (see Supplementary Methods). As the random forest algorithm does not indicate the direction of risk associated with each predictor, this was estimated using univariate logistic regression analyses for each predictor.

#### Model construction and optimisation

To identify the best classification algorithm, seven state-of-the-art machine learning classifiers were implemented: two support vector machines (SVM) (linear and radial bias kernel functions), decision tree, random forest, naive Bayes, multilayer perceptron (MLP), and K-Nearest Neighbours (KNN) (see Supplementary Methods).

Each of the seven machine learning algorithms were initially trained and evaluated on the subset of individuals who had complete data for the predictors selected through RFE. The dataset was split (ratio of 2:1, preserving class proportions) into a training and holdout test set for model development and validation, respectively. Grid search was used to tune the hyperparameters for each model within a five-fold cross-validation, optimizing for its balanced accuracy (see Supplementary Methods, Table E2).

The training dataset was then optimised in an attempt to further improve the performance of the classification algorithms. Multiple imputation using multivariate imputation by chain equations (MICE)^22^, oversampling using an adaptive synthetic sampling approach (ADASYN)^23^, and random under-sampling were implemented in a stepwise approach to address the degree of missing data and class imbalance in the training set (see Supplementary Methods). The seven algorithms were redeveloped, and the hyperparameters were tuned, on each optimised training set in order to identify the best asthma prediction models. As the test dataset was not modified, it acted as a single, unseen complete dataset to evaluate the predictive performance of each developed model.

#### Model evaluation

The predictive performance of each model was evaluated on the test set using measures of discrimination (area under the receiver operating curve (AUC)), sensitivity, specificity, positive and negative predictive values (PPV and NPV), positive and negative likelihood ratios (LR+ and LR-) and F_1_-score. The balanced accuracy of each model (which accounts for the prevalence of each class) was also reported. Performance measures were evaluated at the optimal threshold that maximized the Youden’s Index. The Brier score, which measures the mean squared difference between the predicted probability of the outcome and the observed outcome, was also calculated.

The best CAPE and CAPP models were selected based on their discriminative performance in the test set. To calculate 95% confidence intervals for the performance measures in the test and independent validation datasets, 2000 bootstrap samples were used.

### External Validation

The best CAPE and CAPP models were validated in the Manchester Asthma and Allergy Study (MAAS) cohort^24^ (see Supplementary Methods). Data extracted from MAAS was closely matched to maximise the similarity of predictor and outcome definitions used in the development cohort (Table E3). The prediction outcome of school-age asthma was evaluated at both eight and eleven years.

Model generalisability was assessed in MAAS among three risk groups: i) all individuals (unselected population); ii) individuals with at least one parent with allergic disease (asthma, eczema or allergic rhinitis); and iii) individuals with two parents with allergic disease.

### Sensitivity Analyses

The robustness of the CAPE and CAPP models was evaluated using an alternative definition of school-age asthma that incorporated an objective outcome measure. Using this alternative asthma definition, a child was considered asthmatic if they presented with wheeze in the last 12 months and had bronchial hyper-responsiveness (BHR), defined as a 20% reduction in forced expiratory volume in one second (FEV_1_) following a methacholine challenge test^25^.

Additionally, the resolution of the asthma predictions to distinguish between individuals presenting with distinct wheeze phenotypes throughout childhood and adolescence was assessed. The identification and assignment of individuals in the IOWBC and MAAS into one of five distinct wheeze phenotypes through a latent class analysis has been previously described^26^ (see Supplementary Methods).

### Comparison with regression-based methods

To evaluate the hypothesis that machine learning methods may offer greater predictions of childhood asthma than regression-based methods, the CAPE and CAPP machine learning models were directly compared with equivalent logistic regression models developed using the same predictors.

Where data was available in the IOWBC, the performance of the machine learning models were also compared against their existing regression-based benchmark models, and differences in predictions were evaluated using reclassification tables and net reclassification indices (NRI) (see Supplementary Methods).

### Software

Data cleaning, pre-processing and imputation were performed using R statistical programming language (version 3.5.1)^27^. All other data manipulation and modelling was performed using Python scripting language (version 3.6.8) and the machine learning libraries Scikit-learn^28^ and imbalanced-learn^29^.

## RESULTS

In the IOWBC, 1368 enrolled participants had a defined asthma outcome at age 10, of whom 201 (14.69%) were classified as asthmatic. Asthmatic children at age 10 were significantly more likely to be male, have a lower birthweight, be atopic and experience a range of asthma-like symptoms, both in early life and at preschool age (Table E4). Baseline characteristics between subjects with complete data used for feature selection were largely comparable with the full IOWBC dataset (Table E4).

### Childhood Asthma Prediction in Early-life (CAPE) Model

Data on 39 of the 54 candidate predictors were collected by age two. Complete data on all 39 predictors was available for 490 individuals. RFE identified a subset of eight predictors for inclusion in the CAPE model, with an average balanced accuracy of 64.49% (Table 1). Table E5 details the direction of asthma risk for each selected predictor. Complete data for these eight predictors was available for 765 individuals; 510 (68 asthmatics) and 255 (34 asthmatics) subjects were allocated to the initial training and test sets, respectively. A SVM with a RBF kernel was the best performing classification algorithm for the CAPE model (Table 2A), with moderate predictive performance (AUC=0.71 and Brier score=0.21).

**Table 1.** Optimal subset of predictors selected by RFE for inclusion in the CAPE and CAPP models

| **Table 1. Predictors selected for inclusion in the model (n)** |  | **Average balanced accuracy (%)** |
| --- | --- | --- |
| **CAPE** | (8) - Maternal age, birthweight, total breastfeeding duration, age of solid food introduction, BMI at 1 year, wheeze in early life, cough in early life and maternal socioeconomic status | 64.49 |
| **CAPP** | (12) - Maternal age, birthweight, total breastfeeding duration, age of solid food introduction, BMI at 1 year, BMI at 4 years, wheeze, cough, nocturnal symptoms, atopy status, polysensitisation and maternal socioeconomic status | 74.93 |

**Table 2.**
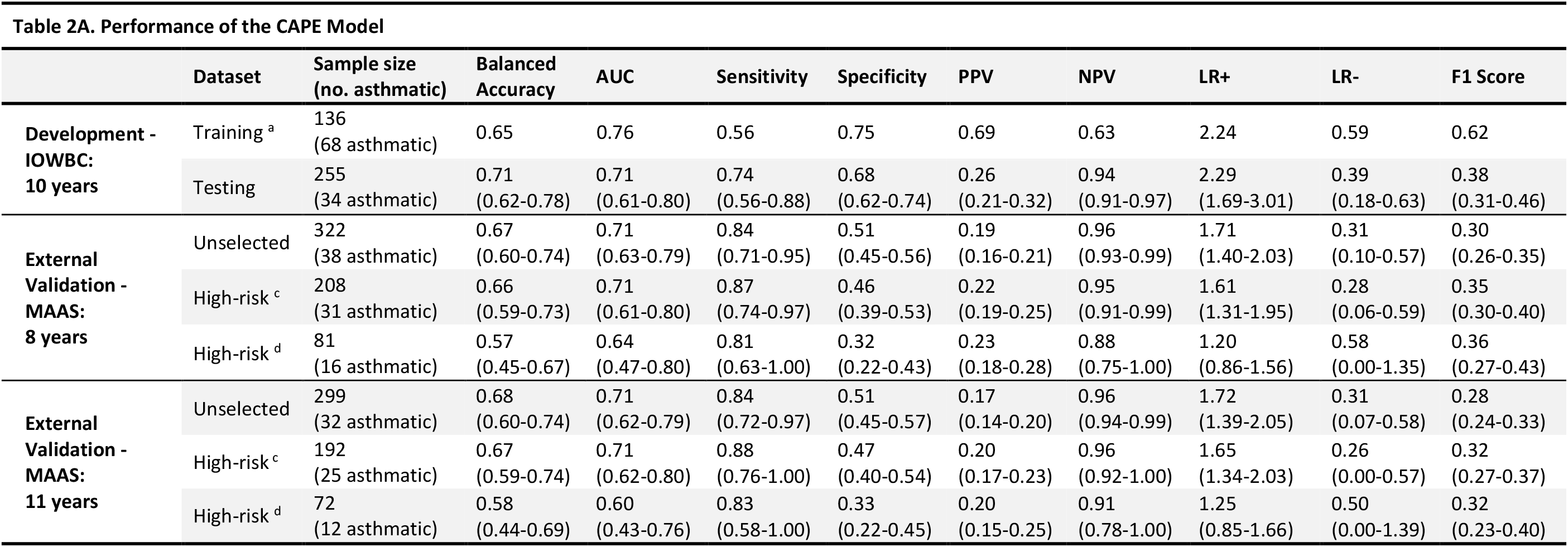
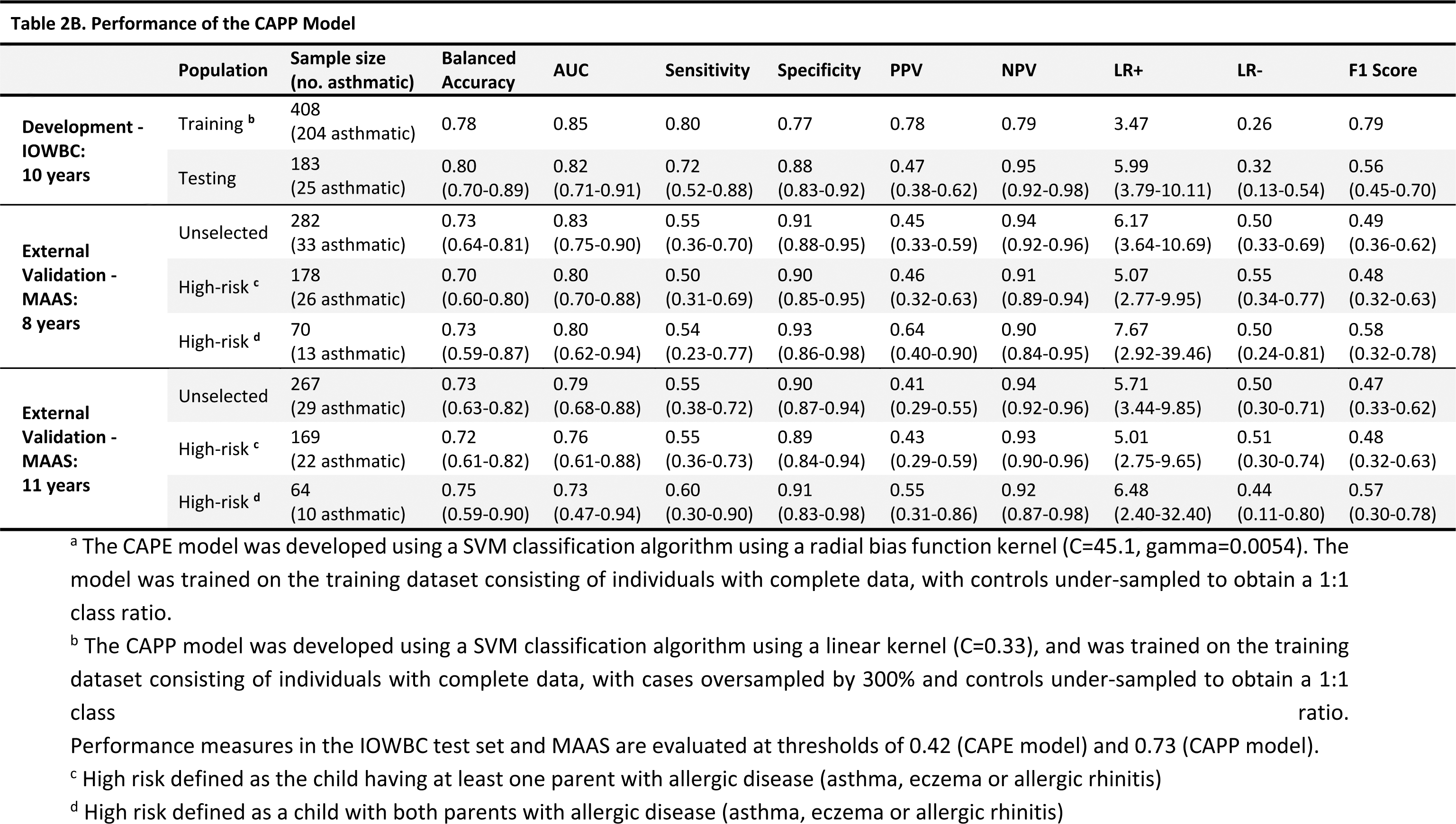
Performance of the CAPE and CAPP models

#### External Validation of the CAPE Model

To predict the development of asthma at the 8-year and 11-year time-points in MAAS, complete data on the eight CAPE predictors was available for 322 and 299 individuals, respectively. Table E6 compares the distribution of predictors in the IOWBC and MAAS. The CAPE model demonstrated moderate generalisability, with a reduction in PPV despite maintaining an AUC=0.71 in the validation study at both 8 and 11 years (Table 2A, Figure 2). In the high-risk subgroups, despite a slight increase in PPV, overall predictive performance decreased (Table 2A).

**Figure 1:**
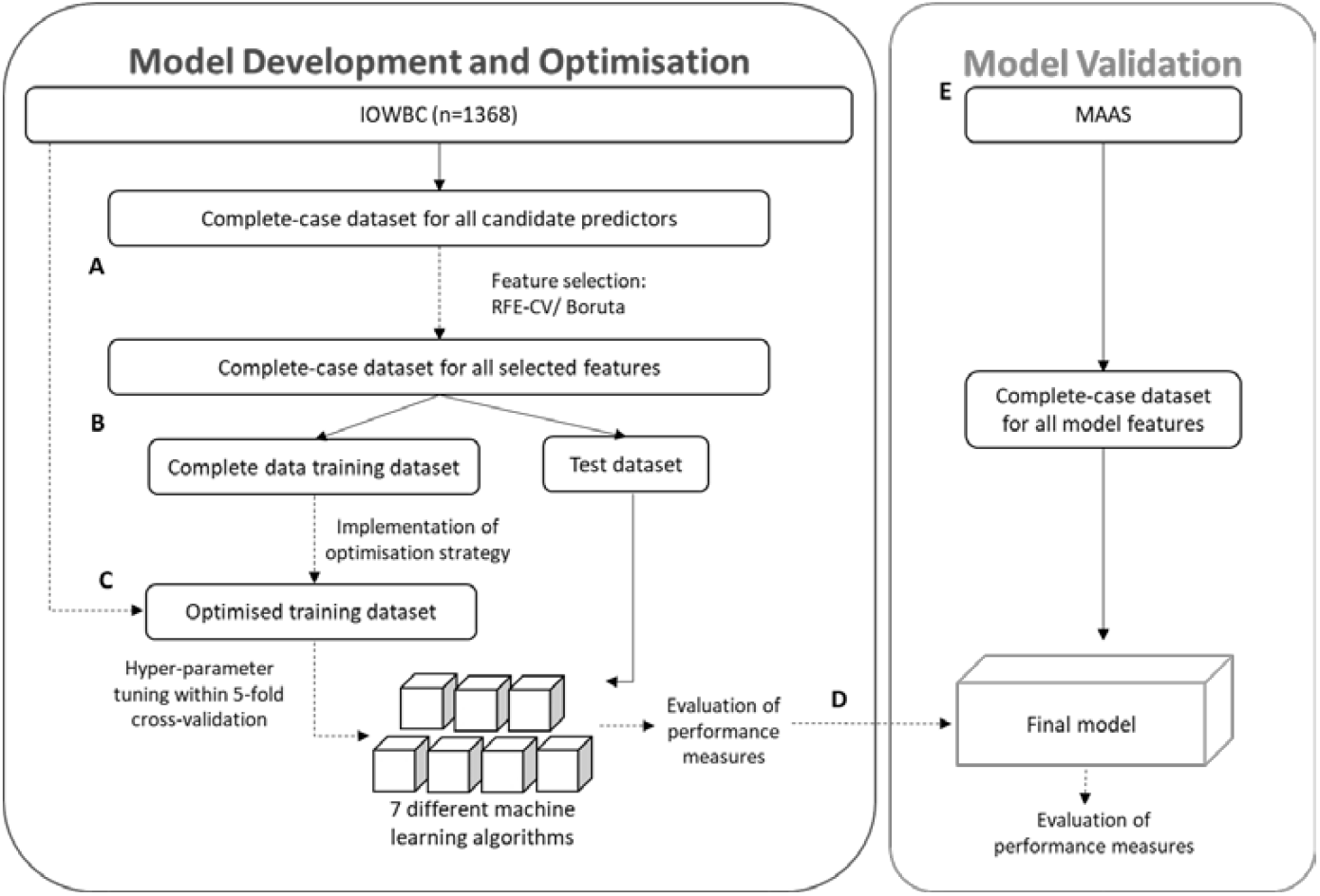
Workflow for the development and validation of asthma prediction models using machine learning approaches. Model development was performed using data from the Isle of Wight Birth Cohort (IOWBC). The following process was performed independently for the construction of the CAPE and CAPP tools. (A) Feature selection was performed using only individuals with complete data for all available candidate predictors. (B-C) Seven machine learning classifiers (two support vector machines with different kernel functions (linear and radial bias function), naïve Bayes classifier, decision tree, multilayer perceptron, random forest and K-nearest neighbours) were developed. Models were first developed using only individuals with complete data for the subset of features identified from feature selection (B), and subsequently redeveloped using optimised training datasets (C). Optimisation of the training dataset consisted of the step-wise application of imputation and resampling (oversampling using ADASYN and random undersampling) to the dataset of all individuals in the IOWBC not allocated to the test dataset, including those with missing predictor data. (D) The best models for use in early life (CAPE tool) and at preschool age (CAPP tool) were selected based on their performance in the test set. (E) The selected models were externally validated in an independent population (Manchester Asthma and Allergy Study, MAAS).

**Figure 2:**
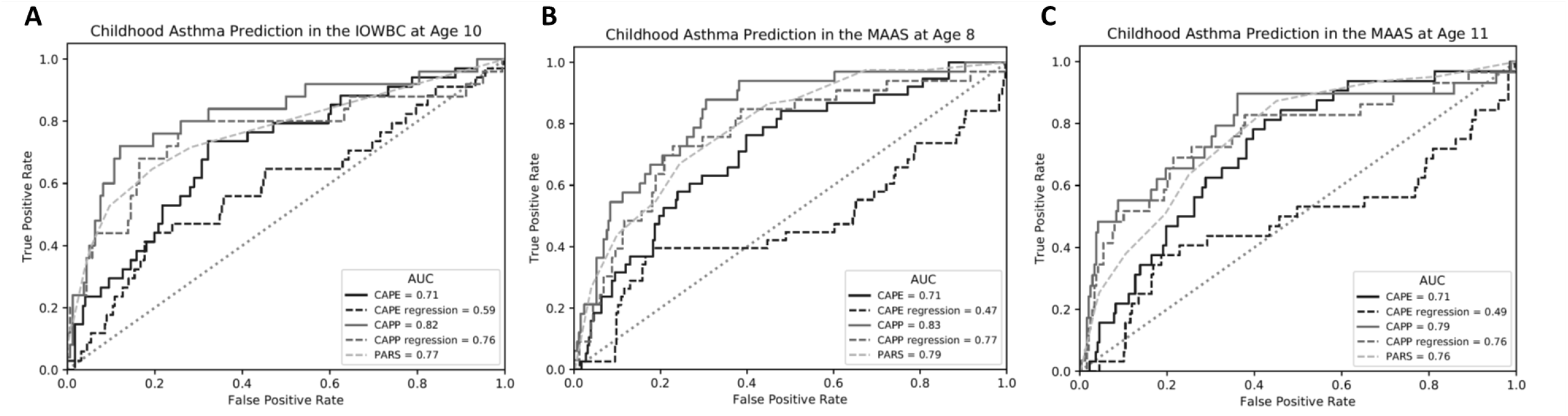
Discriminative performance of the CAPE and CAPP machine learning models, compared against their equivalent regression models and the PARS model. ROC curves comparing the generalisability of the five models in the IOWBC at age 10 (A) and upon validation in MAAS at age 8 years (B) and 11 years (C).

### Childhood Asthma Prediction at Preschool-age (CAPP) Model

For the CAPP model, 373 individuals had complete data for all 54 candidate predictors available by age four. RFE identified an optimal subset of 12 predictors for inclusion in the model, with an average balanced accuracy of 74.93% (Table 1) and direction of asthma risk for each predictor detailed in Table E5. Complete data for these 12 predictors was available for 548 individuals, of whom 365 (51 asthmatics) and 183 (25 asthmatics) individuals were assigned to the initial training and test sets, respectively. The best performing classification algorithm for the CAPP model was a SVM with a linear kernel (Table 2B). The model demonstrated good predictive performance, with AUC=0.82 and Brier score=0.18.

#### External Validation of the CAPP Model

For validation of the CAPP model in MAAS at the 8-year and 11-year time-points, complete data for the 12 CAPP predictors was available for 282 and 267 individuals, respectively. The model demonstrated good generalisability in predicting asthma at both 8 and 11 years (AUC=0.83 and 0.79, respectively) in the unselected MAAS subgroup (Table 2B). PPV remained comparable upon validation (PPV=0.45 and 0.41, respectively), with further improvements reported in both high-risk subgroup validations at both time-points (Table 2B).

##### Sensitivity analysis

Asthma status, based on the alternative asthma definition, was available for 1312 of the 1368 individuals analysed in the IOWBC (prevalence 8.61%). Despite an overall 92.3% agreement, there was a statistically significant difference between the two asthma definitions (p<0.01). This stemmed from a 97.6% agreement for labelling non-asthmatics but only a 53.8% agreement for labelling asthmatics (Figure E1).

A labelled asthma status using the alternative asthma definition incorporating BHR was available for 248 out of 255 individuals in the CAPE test dataset (20 asthmatic) and 179 out of 183 individuals in the CAPP test dataset (18 asthmatic). The CAPE and CAPP models were less robust to predict the alternative asthma outcome (CAPE AUC=0.67 vs 0.71 and CAPP AUC=0.79 vs 0.82). Both models demonstrated an increased sensitivity to predict asthmatics, but the corresponding increase in false positive predictions resulted in PPV reducing by approximately 50% (Table E7).

In the CAPE and CAPP test datasets, 213 and 167 individuals had a defined wheeze phenotype, respectively. For both models, individuals predicted as non-asthmatic at age 10 largely presented with the never/infrequent or late-onset wheeze phenotypes (Figure 3). Both models showed excellent power to predict the persistent wheeze phenotype, with 100% and 90% of individuals with persistent wheeze offered a positive prediction by the CAPE and CAPP models, respectively. Upon validation, the CAPE and CAPP models were able to positively predict 91% and 57% of persistent wheezers, respectively.

**Figure 3:**
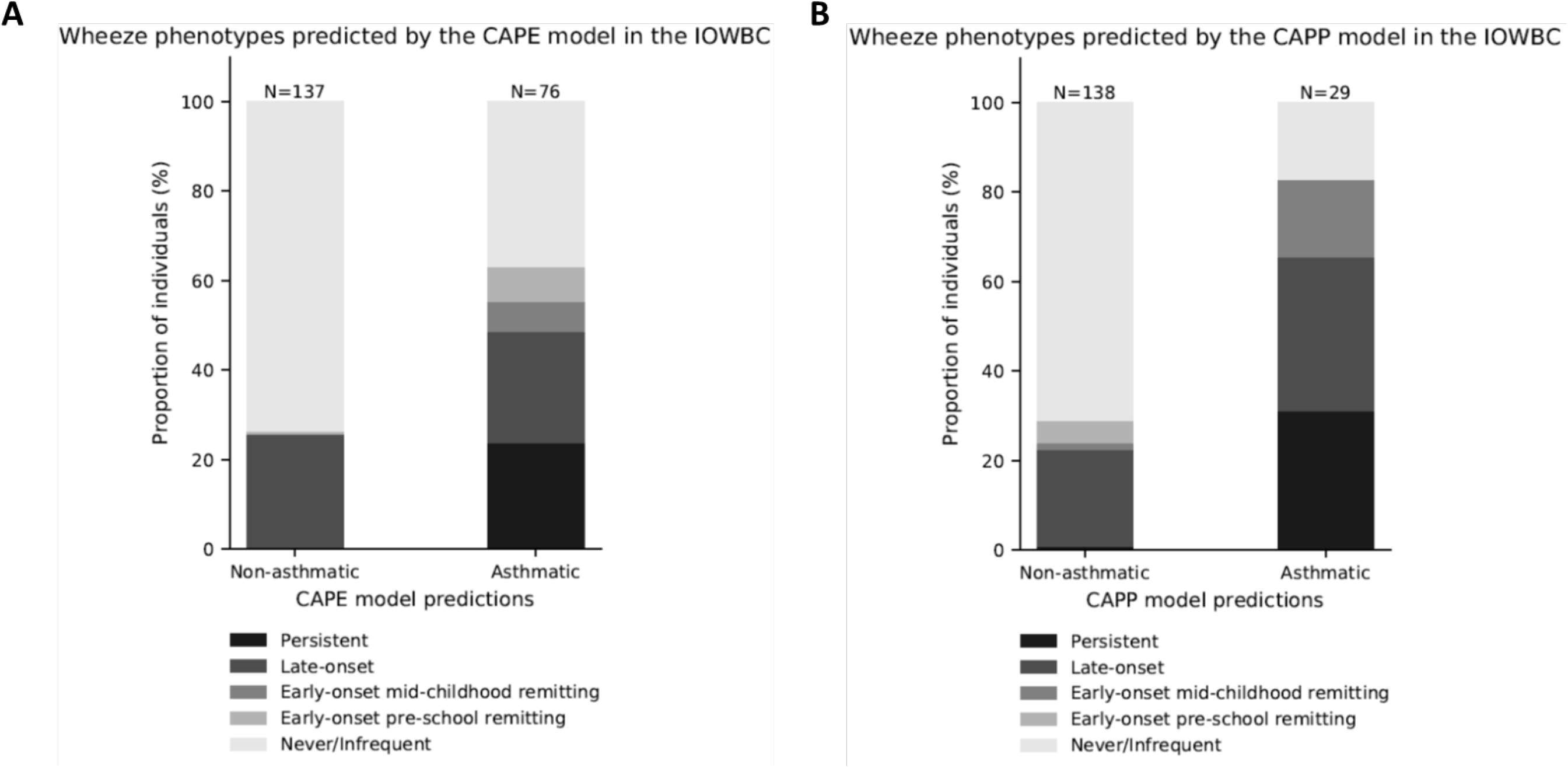
CAPE and CAPP model predictions and corresponding wheeze trajectories. The proportion of individuals corresponding to their most probable wheeze phenotypes is presented for those offered a negative or positive prediction by either the CAPE (A) or CAPP model (B) in the IOWBC (predicted non-asthmatic or asthmatic at age 10, respectively).

##### Comparison with regression-based methods

Both the CAPE and CAPP machine learning models outperformed their equivalent logistic regression models (Table 3, Figure 2). There was a substantial decline in predictive performance of the CAPE model when using a logistic regression model (AUC=0.59 vs 0.71), with predictions being no better than chance in MAAS at 8 and 11 years (AUC=0.47 and 0.49, respectively). Yet, PPV remained comparable with the CAPE machine learning model (PPV=0.25 vs 0.26). Predictive power of the CAPP model also reduced when using a logistic regression model (AUC=0.76 vs 0.82), with a 14% decline in PPV (PPV=0.33 vs 0.47). The CAPP equivalent logistic regression model demonstrated poor generalisability, with a further 11-13% decline in PPV in MAAS; the CAPP machine learning model demonstrated good generalisability upon validation, maintaining similar PPV to that observed in the developmental population.

**Table 3.**
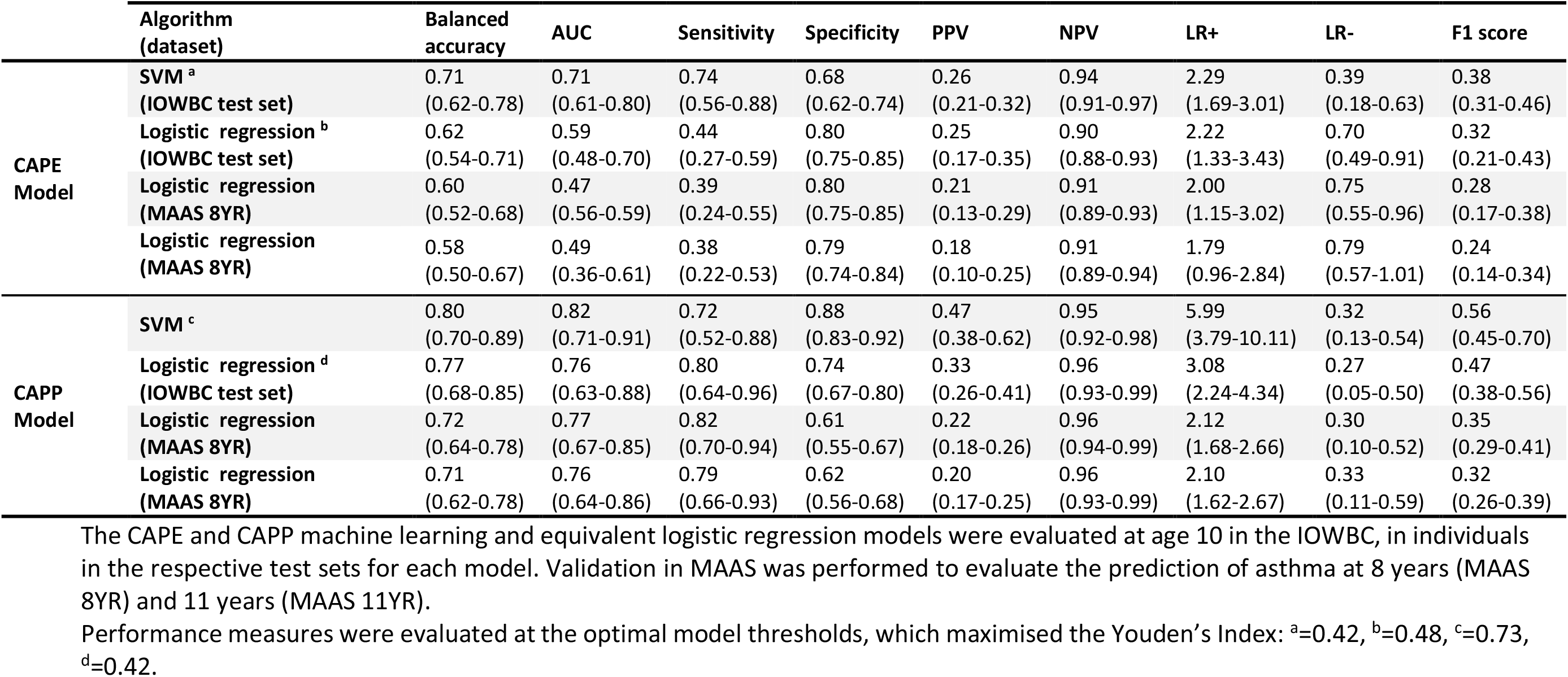
Comparison of the prediction models developed using machine learning and traditional logistic regression algorithms

Of the existing validated regression-based models, the PAPS (Persistent Asthma Predictive Score)^30^ and PARS (Paediatric Asthma Risk Score)^31^ were considered the best performing models comparable with the CAPE and CAPP models, offering predictions in early life and at preschool age, respectively. The well-known Asthma Predictive Index was not replicable due to the absence of eosinophil data in the IOWBC. Similarly, PAPS was unable to be replicated as RAST tests were not performed in the IOWBC. However, data was available to replicate the PARS model (AUC IOWBC=0.77, MAAS 8YR=0.79, MAAS 11YR=0.76). In individuals with predictions available from both the CAPP and PARS models in the IOWBC CAPP test set (n=174), 32% of asthmatic children at age 10 were correctly reclassified as asthmatic by the CAPP model whilst only 5% of non-asthmatics were incorrectly reclassified as asthmatic compared to predictions made by the PARS model (Table 4). Prediction classifications made by the CAPP model remained superior compared to PARS upon validation in MAAS (Table E8).

**Table 4.** Reclassification table showing changes in prediction categorisation between the PARS and CAPP model

|  | Predicted risk (CAPP model) |  |  | Reclassified by CAPP (%) |  |  |  |
| --- | --- | --- | --- | --- | --- | --- | --- |
| Predicted risk (PARS model) | No asthma | Asthma | Total | Increased risk | Decreased risk | Correctly reclassified | NRI |
| No asthma at age 10 (n=149) |  |  |  |  |  |  |  |
| No asthma | 130 | 9<sup>b</sup> | 139 |  |  |  |  |
| Asthma | 1<sup>a</sup> | 9 | 10 | 9(6%) | 1(<1%) | 1(<1%) | -0.05 |
| Total | 131 | 18 | 149 |  |  |  |  |
| Asthma at age 10 (n=25) |  |  |  |  |  |  |  |
| No asthma | 7 | 8<sup>a</sup> | 15 |  |  |  |  |
| Asthma | 0<sup>b</sup> | 10 | 10 | 8(32%) | 0(0%) | 8(32%) | 0.32 |
| Total | 7 | 18 | 25 |  |  |  |  |
|  |  |  | Total | 17 | 1 | 9 |  |

Reclassification table comparing the change in individual asthma predictions with the CAPP model instead of the PARS model (reference model). For the PARS model, categorisations of predictions as asthmatic and non-asthmatic was based on the optimal threshold (cutoff=7) as defined in their original publication. Results are presented separately for individuals who were asthmatic and non-asthmatic at age 10. Values in bold identify the number of individuals who were reclassified into a more appropriate (<sup>a</sup>) or less appropriate (<sup>b</sup>) risk group by the CAPP model with respect to the risk classifications made by the PARS model. NRI=net reclassification index is given separately for true asthmatics and non-asthmatics.

## DISCUSSION

### Summary of findings

Two models predicting school-age asthma at age 10 within a general population were developed using machine learning classification methods. The CAPE model uses a RBF SVM classifier and data from eight predictors to predict school-age asthma in early life. The CAPP model uses a linear SVM classifier and information on twelve predictors available by age four. Both machine learning models offered superior predictive power and generalisability upon external validation compared to equivalent models developed using logistic regression methods as well as existing regression-based models. Whilst the primary prediction outcome was school-age asthma, both models demonstrated greater sensitivity in predicting individuals likely to experience persistent wheeze throughout childhood.

### Comparisons with Existing Models

To date, twenty-one regression-based prediction models have been developed for childhood asthma (reviewed in Kothalawala et al.^7^), of which only six have been externally validated. Using machine learning approaches, both the CAPE and CAPP models were able to surpass the performance of their respective benchmark models based on AUC (CAPE=0.71 vs PAPS=0.66; and CAPP=0.82 vs PARS=0.80). The CAPP model further outperformed all six validated regression-based models based on discrimination (Table E9).

Similar to the CAPE and CAPP models, most published asthma prediction models are very good at ruling out asthma rather than ruling in asthma, resulting partly from low power due to low asthma prevalence^7^. Even if existing models offer good PPV, this often degrades upon validation^7^. Arguably, the need for exploring novel methodologies for asthma prediction is to improve the ability to rule in asthma and improve model generalisability. Indeed, despite having similar asthma prevalence to existing studies, the machine learning-based CAPP model was able to offer a 30% improvement in sensitivity compared to the loose API (sensitivity: CAPP=0.72, API=0.42) and further 10% improvement in PPV compared to its benchmark model, PARS. Importantly, the CAPP machine learning model was more generalisable and able to retain its positive predictive power upon validation compared to its equivalent logistic regression model. Furthermore, direct comparison between the classifications made by the CAPP and PARS models highlighted the improved power for the CAPP model to correctly predict future asthmatics. The moderate but limited predictive power of the CAPE model compared to the CAPP model was unsurprising given the known difficulty of predicting childhood asthma in the first few years of life^32^.

### Predictor selection and availability

Both the CAPE and CAPP models include data collected across multiple time-points -at birth, in early life and at preschool-age (the latter for the CAPP model only) (Figure E2 and E3). Given the changeable nature of asthma development and risk throughout early childhood, the consideration of predictors across multiple time-points allowed the identification of a novel combination of predictors that together helped to improve current asthma predictions. Whilst data collected across multiple time-points may hinder the utility of the model, the required predictor data are typically reported during a child’s routine health visits or tracked in child health records, such as those provided for each newborn child in the UK. Only the predictors of atopy and polysensitisation, which require a skin prick test (SPT) to be performed, may hinder the applicability of the CAPP model. However, the exclusion of these predictors resulted in an 8% reduction in model sensitivity (Table E10); hence, the predictive benefit offered by the inclusion of sensitisation was deemed to outweigh the potential loss in model applicability.

Of the predictors selected for inclusion in the two models, some were well-established risk factors with a clear direction of asthma risk (Table E5). Others were novel predictors (maternal age at the time of the child’s birth, age of solid food introduction and total breastfeeding duration) that did not indicate a clear direction of asthma risk based on univariate logistic regression analysis. The selection of these novel predictors, over other predictors that have more established biological relevance in the literature (such as parental asthma, eczema or allergic rhinitis), may be cautiously accepted by the clinical community. However, the aim of RFE is to identify the subset of features that collectively offer the best predictive accuracy rather than to devise a comprehensive list of risk factors of childhood asthma, which may be biologically sound but lacking in predictive power^33^. Therefore, the inclusion of these novel predictors may be indicative of the improved power for feature selection using machine learning methods to account for the relatedness between features, and uncover previously unidentified predictors, compared to traditional univariate or multivariate regression-based approaches. These predictors could also be surrogates for more biologically relevant features. It is also important to acknowledge the possibility that the selection of these novel predictors may stem from an inherent bias of the random forest algorithm to assign greater importance to features which are continuous or which have a large number of categories^34^. However, as the CAPE and CAPP models developed using these selected predictors demonstrated improved performance against existing prediction models, any bias stemming from the feature selection process did not appear to hinder the selection of features that were truly predictive of school-age asthma.

### Prediction generalisability, robustness and resolution

In the unselected MAAS cohort, the CAPE and CAPP models showed moderate-good generalisability to predict asthma across school ages (age 8 and 11), despite the marginal decline in the PPV of the CAPE model. Validation in high-risk MAAS subgroups showed the PPV of both models to increase with the number of allergic parents, suggesting that confidence in ruling in asthma improves in high-risk groups; but replication in a larger study population is required.

In addition, the lack of a clear definition for asthma is an unavoidable limitation in epidemiological studies^35^. The asthma definition used in this study aimed to account for children with a clinical indication of asthma (physician diagnosed) who were actively symptomatic, but also those potentially asymptomatic at the time of assessment due to the use of symptom relieving medications. It is likely that the majority of children in the IOWBC with clinically relevant asthma were detected. Whilst the CAPE and CAPP models were robust in predicting non-asthmatics using an alternative asthma definition of wheeze and BHR, they had reduced power to predict true asthmatics (∼50% decline in PPV). The latter may be explained by objective tests such as spirometry and BHR being influenced by treatment; potential asthmatics on controller medications, whom the models are trained to identify as asthmatic, may be considered as non-asthmatic with the alternative definition, resulting in greater false positive predictions.

Evidently, asthmatics, by either definition, identify a highly heterogeneous group; better definitions or explorations of asthma sub-phenotypes are warranted. As the aim of this study was to compare whether machine learning approaches could improve upon existing regression-based models that predict childhood asthma, the primary prediction outcome for this study was restricted to school-age asthma rather than predicting asthma phenotypes. However, acknowledging the importance of exploring specific sub-phenotypes of asthma, the resolution of the machine learning models to inform on an individual’s future wheeze trajectory was also explored. Notably, both the CAPE and CAPP models showed excellent sensitivity to predict individuals with a persistent wheeze phenotype; these individuals would likely benefit from early primary or secondary asthma prevention/ management.

### Strengths and limitations

This study had a number of strengths. First, each model was developed to make timely predictions to identify future asthmatics within a general population, rather than among those whom physicians already consider to be at high-risk (mainly those experiencing wheeze or with a familial history of asthma/allergy). Second, by utilising machine learning methods, which have infrequently been applied for predicting childhood asthma, novel predictors of school-age asthma were identified and the developed models offered improved predictive performance over current regression-based methods, with an improvement in PPV when predicting asthma at preschool age. Third, both models were externally validated and demonstrated good generalisability to predict school-age asthma across multiple time-points, without degrading the predictive power to rule in asthma (particularly with the CAPP model). Finally, the two models displayed moderate resolution to inform on a child’s wheeze trajectory, with excellent sensitivity to predict persistent wheeze.

However, this study was limited by both model development and validation being conducted in predominantly Caucasian populations; validation in ethnically diverse populations is warranted to assess the widespread generalisability of the two models. Machine learning also requires large datasets – the introduction of more data would undoubtedly improve the performance of the machine learning models further. Finally, whilst biological data was available in the IOWBC, only clinical and environmental predictors were considered. It is possible that the consideration of such predictors might significantly improve childhood asthma predictions further; however, the aim of this study was to explore whether machine learning methods could surpass the predictive ceiling that existing logistic regression methods appeared to be limited to. Hence, to provide a fair comparison with existing regression-based models, asthma biomarkers were not incorporated into this study.

### Conclusion and Future Work

Using machine learning approaches, the CAPE and CAPP models were able to surpass the predictive performance of similar models developed using regression-based methods. Whilst both models were generalisable in an independent population, the CAPP model also demonstrated superior predictive power to rule in true asthmatics compared to its benchmark model, which was retained upon validation. Both models also demonstrated excellent sensitivity to predict a subgroup of persistent wheezers. Therefore, rather than developing an all-encompassing asthma prediction tool, further research into predicting specific ‘asthmas’ using machine learning approaches may offer greater predictive insight. Finally, continued exploration of machine learning approaches and the identification and integration of novel biomarkers is warranted to further improve the power to predict future childhood asthma.

## Data Availability

Access to data from the Isle of Wight Birth Cohort can be made available upon request. Further information can be found at www.allergyresearch.org.uk/ or from: Cohort Profile: The Isle Of Wight Whole Population Birth Cohort (IOWBC). Int J Epidemiol. 2018 Aug 1;47(4):1043-1044i. doi: 10.1093/ije/dyy023. Source code for the development and use of the prediction models can also be made available upon request.

## Abbreviations

AUC: Area under the receiver operating curve
BHR: Bronchial hyper-responsiveness
RFE: Recursive feature elimination
LR-: Negative likelihood ratio
LR+: Positive likelihood ratio
NPV: Negative predictive value
PPV: Positive predictive value
SPT: Skin prick test

## Acknowledgments

The authors would like to acknowledge the help of all the staff at the David Hide Asthma and Allergy Research Centre in undertaking the assessments of the Isle of Wight birth cohort. The authors would also like to thank the IOWBC and MAAS study participants and their parents for their continued support and enthusiasm. Recruitment and initial assessment for the first 4 years of age for the IOWBC was supported by the Isle of Wight Health Authority. The 10-year follow-up of the IOWBC was funded by the National Asthma Campaign, UK (Grant No 364). MAAS was supported by the Asthma UK Grants No 301 (1995-1998), No 362 (1998-2001), No 01/012 (2001-2004), No 04/014 (2004-2007), BMA James Trust (2005) and The JP Moulton Charitable Foundation (2004-current), The North west Lung Centre Charity (1997-current) and the Medical Research Council (MRC) G0601361 (2007-2012), MR/K002449/1 (2013-2014) and MR/L012693/1 (2014-2018). UNICORN (Unified Cohorts Research Network): Disaggregating asthma MR/S025340/1.

## STELAR/UNICORN Investigators

Graham C Roberts, Human Development and Health, Faculty of Medicine, University of Southampton, David Hide Asthma and Allergy Research Centre, Isle of Wight and NIHR Southampton Biomedical Research Centre, University Hospitals Southampton NHS Foundation Trust, Southampton, UK

Steve W Turner, Child Health, University of Aberdeen, Aberdeen, UK

Raquel Granell, MRC Integrative Epidemiology Unit, Population Health Sciences, Bristol Medical School, University of Bristol, UK

Sadia Haider, National Heart and Lung Institute, Imperial College London, UK

Sara Fontanella, National Heart and Lung Institute, Imperial College London, UK

Paul Cullinan, National Heart and Lung Institute, Imperial College London, UK

