## Supplementary Material for "Development of Childhood Asthma Prediction Models using Machine Learning Approaches"

### Methods

#### Datasets

**Developmental Study Population: Isle of Wight Birth Cohort**

Of the 1536 newborns born between January 1989 and February 1990, 1456 were recruited and prospectively followed up at 1, 2, 4, 10, 18 and 26 years of age[E1]. Individuals lost to follow-up were significantly more likely to have asthmatic siblings, be of low birthweight and have parents who smoked. At each visit, information was collected on clinical symptoms of asthma and other allergic diseases as well as lifestyle and environmental exposures through hospital records, physical examinations and study-specific parent/participant questionnaires. Skin prick testing (SPT) was performed in infants with allergy-related symptoms at the 1-year and 2-year follow-ups, and in all participants from the 4-year follow-up onwards.

**Validation Study Population: Manchester Asthma and Allergy Study**

The MAAS cohort is a longitudinal, whole-population UK birth cohort established in 1995 to study the development of asthma and other atopic disorders in childhood[E2]. In brief, parents were recruited between 1995 and 1997, and 1184 children followed up at 1, 3, 5, 8, 11, 13-16 and 18 years. Medical records and validated questionnaires were used to collect data on clinical symptoms of allergy and asthma and environmental exposures. Blood samples were taken and SPT and lung function tests were performed from the three-year follow-up onwards.

Early life predictor data was collected from the 1-year and 3-year follow-ups in MAAS (1-year and 2-year follow-ups in the IOWBC), and from the 5-year follow-up for preschool predictors (4-year follow-up in the IOWBC).

#### Feature Selection

**Pre-processing of candidate predictors**

Pre-processing of all candidate predictor data included the removal of extreme outliers (±4SD) present in each continuous variable and one-hot encoding of nominal variables into separate binary variables. Child BMI reported at 1 and 4 years old were standardised against the British 1990 growth reference[E3].

**Recursive Feature Elimination**

Feature selection was conducted using Recursive Feature Elimination (RFE) with a random forest algorithm. RFE initially builds a random forest classifier using all of the candidate features and evaluates its predictive performance[E4,5]. In an iterative process, the lowest ranking feature, in terms of gini feature importance, is removed and the random forest classifier is rebuilt on the remaining feature subset and the predictive performance of the classifier is re-evaluated. This process was performed within a five-fold cross-validation framework whereby: i) the dataset was split into five equal folds, preserving the number of asthma cases in each fold; ii) the random forest classifier for RFE was built on 4 of the 5 folds and the balanced accuracy of predicting both asthmatic cases and non-asthmatic controls was evaluated on the remaining fold; iii) the optimal subset of predictors was identified based on the random forest model that achieved the highest cross-validation balanced accuracy score. A variation of the random forest algorithm (balanced random forest), which randomly under-samples the majority class in each bootstrap, was used to account for the class imbalance present in the dataset. For feature selection, all continuous variables were standardised to zero mean and unit variance.

**Assessing the directionality of selected predictors**

The random forest algorithm used in RFE is able to select predictors based on their predictive importance, however it is unable to offer insight into the directionality of each predictor. As a result, univariate logistic regression was performed on each predictor to evaluate whether they incurred a risk or protective effect on the development of school-age asthma.

#### Classification Algorithms

**Support Vector Machine**

Support vector machines aim to construct a separating hyperplane between the outcome classes. A soft margin is used to penalize misclassifications whilst trying to optimise the best classification. When dealing with high-dimensional data, SVM exploits the kernel trick to map the data onto a higher dimensional space in order to construct the separating hyperplane between the outcome classes[E6,7]. This study utilized three different kernel functions - linear, radial basis function (RBF), and polynomial - for constructing the best support vector classifier were used.

**Decision Tree**

Decision trees aim to stratify the predictor space using simple splitting rules. Starting at the top (root node) of the tree, the predictor space is stratified at each internal node. Predictor variables which create the best separation of the outcome classes are calculated at each node, with better splitting variables utilised at nodes higher in the tree structure. The final nodes at the bottom of the tree, at which no further separations are made, are referred to as leaves or terminal nodes and provide the final classification[E6].

**Random Forest**

An ensemble classifier that aggregates the decisions of multiple decision trees. For the development of each tree, a bootstrapped dataset the same size as the original dataset is created by randomly resampling the original dataset. Unlike the decision tree algorithm, each tree in a random forest only considers a random subset of variables for stratifying the predictor space. As a result, the correlation between the trees will be reduced. Although each tree will have low bias and high variance, the process of bootstrapping and aggregating decisions across the trees to make a final classification (known as bagging), will result in predictions of low variance and high accuracy[E6].

**Naïve Bayes**

Based on conditional probability, this algorithm is underpinned by the assumption that each feature is independent of the other in determining the outcome class. Implementation of the Naïve Bayes algorithm requires assumptions of each class' prior probability distribution, commonly Gaussian and multinomial/Bernoulli distribution for continuous and categorical features, respectively[E6].

**Multilayer Perceptron**

A simple feed-forward artificial neural network which is well-suited to distinguish between non-linearly separable data through a network of nodes. A MLP consists of a minimum of three layers; an input layer, at least one hidden layer and an output layer. Each neuron in the input layer is fully connected to each neuron in the next layer. These connections are weighted, with the weights optimised through a process of backpropagation which aims to minimize the error of the output layer which determines the class classification[E8,9].

**K-Nearest Neighbours**

KNN is an instance-based learning algorithm which utilises the feature space to classify unknown data points based on a number of data points in close proximity (nearest neighbours) for which the class labels are known. Based on the conditional probability of the unknown data point belonging to each class, a final classification is made[E6].

**Pre-processing of the training and test set**

Prior to model development (and redevelopment), the continuous variables in the training dataset were standardized to a mean of zero and unit variance, and the same standardisation properties were applied to the test set.

**Hyperparameter tuning**

The hyperparameters of each machine learning algorithm were tuned during the training process to optimise for the model's balanced accuracy. Balanced accuracy was chosen as the optimisation metric in order to maximize the predictive accuracy of both outcome classes whilst accounting for the inherent class imbalance of the training dataset.

To identify the optimal hyperparameters, a grid search was used to systematically search over a range of each model's hyperparameters within a 5-fold cross-validation (ranges detailed in Table E2). To reduce computational time, a random search was performed for the SVM algorithms to narrow the hyperparameter ranges specified for evaluation in the grid search. The Naïve Bayes algorithm did not require any hyperparameter tuning (Table E2).

#### Optimisation of the training dataset

Due to potential bias of using only complete data for model training, the training set was optimised in an attempt to further improve the predictive performance of the classification algorithms.

First, missing predictor data for all individuals that were not allocated to the test set, (n=1185 and n=1119 for the CAPE and CAPP models, respectively), were imputed using Multivariate Imputation by Chain Equation (MICE). ADASYN (an adaptive synthetic sampling approach) was then applied to the imputed training dataset to generate new examples of the minority class (asthmatic cases), with a bias towards increasing the number of difficult to classify examples. The effect of oversampling was assessed across a range of levels – the number of asthma cases were increased by 0%, 25%, 50%, 100%, 150%, 200%, 250% and 300%. Finally, random under-sampling was applied to each of the imputed and oversampled training datasets in order to completely balance the number of asthmatic cases and non-asthmatic controls (1:1 ratio).

**Multivariate Imputation by Chain Equation (MICE)**

Multivariate imputation by chain equation is a type of multiple imputation used under the assumption that data is MAR. By performing multiple impitations and generating a set of plausible estimates, MICE aims to account for the statistical uncertainty associated with the imputation[E10]. MICE can be applied to datasets containing variables of mixed datatypes. For each datatype, a different imputation model can be used[E10,11] .

In the implementation of MICE, all missing values are initially assigned a placeholder value based on mean imputation or random sampling (with replacement) of the observed data for each variable. For the first variable with missing data, $x_1$, the placeholder values are removed and $x_1$ is regressed on the remaining variables [$x_2$, $x_3$, …, $x_i$]. The regression is limited to only those examples for which $x_1$ was observed. The missing values for $x_1$ are then predicted from the posterior predictive distribution generated by the imputation model. This process is repeated for the remaining missing variables, where for example, $x_2$ is regressed on the remaining variables ([$x_3$, $x_4$, …, $x_i$] and the newly imputed variable ($x_1$), again, limited to examples with observed data for $x_2$. One cycle is complete when all of the variables with missing data have been imputed. Numerous cycles are performed in order to converge the distribution parameters of each variable and create a single dataset of stable imputation estimates. To generate multiple (*m*) imputed datasets, this entire process is repeated *m* times[E10,11] .

Following the imputation stage and the generation of multiple imputed datasets, subsequent analyses should be conducted on each of the *m*-imputed datasets and the results are pooled. The pooled results provide estimates with confidence intervals, addressing the statistical uncertainty of the imputation[E10,11]. However, due to the need to tune each of the machine learning algorithm to establish a single model with a single set of tuned parameters, a single imputed dataset was required for model development. To form a single imputed dataset, the imputed values generated across the *m* imputed datasets were averaged, with the mean and modal imputed values taken for the continuous and categorical variables, respectively.

**Adaptive Synthetic (ADASYN) sampling**

Adaptive Synthetic (ADASYN) sampling is an example of a synthetic data generation approach which aims to generate examples of the minority class in order to improve imbalances in data[E12]. The approach is based on the KNN algorithm. ADASYN can specify the construction of datasets with varying degrees of balance. In contrast to randomly oversampling examples of the minority class, the synthetic examples generated through ADASYN are informed by a density distribution of weights for examples of the minority class. The weight assigned to each example is determined by the ratio of examples belonging to the minority class in its k-nearest neighbours. These weights correspond to the learning difficulty of each example and subsequently determines the number of synthetic examples of the minority class that needs to be generated. For example, a difficult to classify example of the minority class (i.e. one that is similar to examples of the majority class) will have a small ratio of minority examples within its k nearest neighbours therefore will have a large weight. Subsequently, a greater number of synthetic examples will be generated based on this minority example. As a result, the learning model will have a greater opportunity to learn from difficult to classify examples of the minority class in addition to reducing the bias of the model by correcting for the class imbalance[E12].

**Random Undersampling**

The number of non-asthmatic individuals in the training datasets were randomly undersampled. The training dataset was shuffled and a random subset of non-asthmatic individuals were removed in order to balance the outcomes classes in the training dataset – i.e. 1:1 ratio of asthmatic and non-asthmatics.

#### Sensitivity Analyses

**Assessing the resolution to predict childhood wheeze phenotypes**

A latent class analysis of 7,719 individuals from five UK birth cohorts, including the IOWBC, identified five distinct phenotypes of wheeze - never/infrequent wheeze, early onset preschool remitting, early onset mid-childhood remitting, persistent, and late-onset wheeze (full details on the analysis can be found in [E13]). In the IOWBC, 912 individuals had wheeze data available across the five time-points required to evaluate their wheeze trajectory. Based on the latent class analysis, each individual was provided with a probability for belonging to each wheeze phenotype. In this analysis, individuals were categorised into their most probable wheeze phenotype.

**Comparison between the machine learning models and existing logistic regression models**

The developed machine learning models were compared against current published models. The API, the most widely known asthma prediction tool, was unable to be replicated due to the absence of eosinophil data in the IOWBC. Of the remaining validated regression-based models, the PAPS (Persistent Asthma Predictive Score)[E14] and PARS (Paediatric Asthma Risk Score)[E15] were considered the best performing models comparable with the CAPE and CAPP models, offering predictions in early life and at preschool age, respectively. However, PAPS was also unable to be replicated as RAST tests were not performed in the IOWBC. PARS was able to be replicated in both the IOWBC and MAAS.

For replication of the PARS model in the IOWBC, race was not reported for each individual; reported cohort demographics suggest the cohort is 96% Caucasian, hence all individuals were assumed to be Caucasian in this replication. All individuals with complete data for the PARS predictors and the asthma outcome were included in the analysis (predicting asthma in the IOWBC at age 10: n=913, in MAAS at age 8 years: n=552, in MAAS at age 11 years: n=487). Models were compared against the CAPP machine learning models based on AUC.

Individuals with predictions from both the CAPP and PARS models were used to compare the differences in individual predictions between the two models. In the IOWBC, only individuals in the test set (i.e. not used to train the model) were included. Reclassification tables were used to evaluate the differences in predictions on an individual level, for asthmatics and non-asthmatic individuals separately[E16]. The table presents the differences in prediction categorization using the CAPP model compared to the PARS model. The net proportion of individuals reclassified by the CAPP model to a more appropriate prediction group was summarized using the net reclassification indices for true future asthmatics and non-asthmatics separately (NRIevent and NRInon-event, respectively)[E16]:

$$NRIevent = P(up|event)-P(down|event)$$

$$NRInon\text{-}event = P(down|non\text{-}event)-P(up|non\text{-}event)$$

### Supplementary Material References

E1. Arshad SH, Holloway JW, Karmaus W, et al. Cohort Profile: The Isle Of Wight Whole Population Birth Cohort (IOWBC). International Journal of Epidemiology 2018;47:1043-4i.
E2. Custovic A, Simpson BM, Murray CS, Lowe L, Woodcock A. The National Asthma Campaign Manchester Asthma and Allergy Study. Pediatric Allergy and Immunology : Official Publication of the European Society of Pediatric Allergy and Immunology 2002;13:32-7.
E3. Cole T. The LMS method for constructing normalized growth standards. Eurpoean Jounal of Clinical Nutrition 1990;44:45-60.
E4. Isabelle Guyon JW, Stephen Barnhill. Gene Selection for Cancer Classification using Support Vector Machines. Machine Learning 2002;46:389-422.
E5. Granitto PM, Furlanello C, Biasioli F, Gasperi F. Recursive feature elimination with random forest for PTR-MS analysis of agroindustrial products. Chemometrics and Intelligent Laboratory Systems 2006;83:83-90.
E6. James G, Witten D, Hastie T, Tibshiran R. An Introduction to Statistical Learning. 1 ed: Springer-Verlag New York; 2013.
E7. Ben-Hur A, Ong CS, Sonnenburg S, Scholkopf B, Ratsch G. Support vector machines and kernels for computational biology. PLOS Computational Biology 2008;4:e1000173.
E8. Hastie T, Tibshirani R, Friedman J. The Elements of Statistical Learning: Springer; 2001.
E9. M.W.Gardner SRD. Artificial neural networks (the multilayer perceptron) - A review of applications in the atmospheric sciences. Atmospheric Environment 1998;32:2627-36.
E10. White IR, Royston P, Wood AM. Multiple imputation using chained equations: Issues and guidance for practice. Statistics in Medicine 2011;30:377-99.
E11. Azur MJ, Stuart EA, Frangakis C, Leaf PJ. Multiple imputation by chained equations: what is it and how does it work? International Journal of Methods in Psychiatric Research 2011;20:40-9.
E12. Haibo He YB, Edwardo A. Garcia, and Shutao Li. ADASYN: Adaptive Synthetic Sampling Approach for Imbalanced Learning. IEEE International Joint Conference on Neural Networks (IEEE World Congress on Computational Intelligence). Hong Kong2008:1322-8.
E13. Oksel C, Granell R, Haider S, et al. Distinguishing Wheezing Phenotypes from Infancy to Adolescence. A Pooled Analysis of Five Birth Cohorts. Annals of the American Thoracic Society 2019;16:868-76.
E14. Vial Dupuy A, Amat F, Pereira B, Labbe A, Just J. A Simple Tool to Identify Infants at High Risk of Mild to Severe Childhood Asthma: The Persistent Asthma Predictive Score. Journal of Asthma 2011;48:1015-21.
E15. Biagini Myers JM, Schauberger E, He H, et al. A Pediatric Asthma Risk Score to better predict asthma development in young children. Journal of Allergy and Clinical Immunology 2018;143:1803-10.e2.
E16. Kerr KF, Wang Z, Janes H, McClelland RL, Psaty BM, Pepe MS. Net reclassification indices for evaluating risk prediction instruments: a critical review. Epidemiology 2014;25:114-21.

### Supplementary Tables

Table E1 List of candidate predictors of childhood asthma

| Candidate predictor | Definition |
|---|---|
| **Family History** | |
| **Maternal smoking at birth** | Maternal smoking status during pregnancy |
| **Paternal smoking at birth** | Paternal smoking status during pregnancy |
| **Maternal asthma** | Maternal asthma status |
| **Maternal eczema** | Maternal eczema status |
| **Maternal hay fever** | Maternal hay fever status |
| **Paternal asthma** | Paternal asthma status |
| **Paternal eczema** | Paternal eczema status |
| **Paternal hay fever** | Paternal hay fever status |
| **Parity** | Position of child in the family |
| **SES** | Maternal socioeconomic status |
| **Prenatal/ at birth** | |
| **Maternal age** | Maternal age at pregnancy |
| **Prematurity** | Gestation age |
| **Caesarean delivery** | Child birth through caesarean delivery |
| **Total breastfeeding** | Total breastfeeding duration |
| **Exclusive breastfeeding** | Exclusive breastfeeding duration |
| **Solid food introduction** | Age, in months, at which solid foods were introduced to the child's diet |
| **Birthweight** | Birth weight (kg) |
| **Sex** | Child's gender |
| **Season of birth** | Season of birth |
| **Dog** | Household pet dog during pregnancy |
| **Cat** | Household pet cat during pregnancy |
| **Furry pet** | Household furry pet during pregnancy - dog, cat or other animal |
| **Early life (1 and 2 year follow-up)** | |
| **SDS BMI** | BMI at age 1, standardised against the British 1990 growth reference |
| **Wheeze** | Occurrence of wheezing before age 2 |
| **Wheeze without cold** | Likely occurrence of wheezing in the absence of a cold before age 2 |
| **Cough** | Occurrence of cough before age 2 |
| **Nasal symptoms** | Occurrence of nasal symptoms before age 2 |
| **Chest infection** | Occurrence of chest infections before age 2 |
| **Nocturnal symptoms** | Occurrence of nocturnal asthma symptoms before age 2 |
| **Eczema** | Eczema status by age 2 |
| **Hay fever** | Hay fever status by age 2 |
| **Atopy** | Atopy status (sensitisation to one or more allergens) by age 2 |
| **Monosensitisation** | Sensitisation to one allergen by age 2 |
| **Polysensitisation** | Sensitisation to two or more allergens by age 2 |
| **Parental smoking** | Household parental smoking status by age 2 |
| **Dog** | Household pet dog by age 2 |
| **Cat** | Household pet cat by age 2 |
| **Furry pet** | Household furry pet (dog, cat or other animal) by age 2 |
| **Early-life living on a farm** | Main residence on a farm in the first year of life |

| Candidate predictor | Definition |
| --- | --- |
| **Preschool age (4 year follow-up)** | |
| **BMI** | BMI at age 4, standardised against the British 1990 growth reference |
| **Wheeze** | Occurrence of wheezing at age 4 |
| **Wheeze without cold** | Likely occurrence of wheezing in the absence of a cold at age 4 |
| **Cough** | Occurrence of cough at age 4 |
| **Nasal symptoms** | Occurrence of nasal symptoms at age 4 |
| **Nocturnal symptoms** | Occurrence of nocturnal asthma symptoms at age 4 |
| **Eczema** | Eczema status at age 4 |
| **Hay fever** | Hay fever status at age 4 |
| **Atopy** | Atopy status (sensitisation to one or more allergens) at age 4 |
| **Monosensitisation** | Sensitisation to one allergen at age 4 |
| **Polysensitisation** | Sensitisation to two or more allergens at age 4 |
| **Parental smoking** | Household parental smoking status at age 4 |
| **Dog** | Household pet dog at age 4 |
| **Cat** | Household pet cat at age 4 |
| **Furry pet** | Household furry pet (dog, cat or other animal) at age 4 |

Table E2 Hyperparameter tuning criteria for each of the seven machine learning algorithms

| Algorithm | Hyperparameters | Description | Search range |
|---|---|---|---|
| **Support Vector Machine** | Cost | Regularisation term | 100 values between $10^{-3}$ and $10^{2}$ [a] |
| | Gamma | Scalar tem for the RBF and polynomial kernels | 100 values between $10^{-2}$ and $10^{2}$ [a] |
| | Degree | Degree term for the polynomial kernel | 1,2,3,…,10 |
| **Decision Tree** | Max tree depth | The maximum depth each tree should be constructed to | 1,2,3,…,32 or None |
| | Min samples split | The minimum number of samples needed to split a node | 2,3,4,…,11 |
| | Max features | The maximum number of features to consider to find the best split | 'log2', 'sqrt', None |
| | Splitter | Criteria used to choose the split at a node | 'best', 'random' |
| | Criterion | Criteria used to determine the quality of a node split | Gini, entropy |
| **Random forest** | N estimators (trees) | The number of trees used to construct the forest | 1,2,4,8,16,32,64,100,200 |
| | Max tree depth | The maximum depth each tree should be constructed to | 1,2,3,…,32 |
| | Min samples split | The minimum number of samples needed to split a node | 2,3,4,…,11 |
| | Max features | The maximum number of features to consider to find the best split | 'log2', 'sqrt', None |
| | Criterion | Criteria used to determine the quality of a node split | Gini, entropy |
| | Bootstrap | Determines whether bootstrapping with replacement should be used to build the trees | True, False |
| **Multilayer Perceptron** | Hidden layers | The number of neurons in each hidden layer | (1,),(2,),…(11,) (1,1),(2,2),…(11,11)[b] |
| | Activation | The activation function for the hidden layers | 'relu', 'identity', 'tanh', 'logistic' |
| | Solver | Criteria used to optimise the weights of the connections | 'lbfgs', 'sgd', 'adam' |
| | Alpha | Regularisation term | $10^{-1}$ ,$10^{-2}$, $10^{-6}$ |
| | Learning rate | The rate at which to update the weights | 'constant', 'invscaling', 'adaptive' |
| | Initial learning rate | The initial learning rate | 0.1,0.2,…,0.9 |

| Algorithm | Hyperparameters | Description | Search range |
|---|---|---|---|
| **KNN** | Number of neighbours (k) | The number of neighbours | 1,2,3,…,100 |
| | Weight | Determines whether each neighbour should be weighted equally or based on their distance | Uniform, distance |
| | Power | Specifies the distance measure to use | Manhattan, Euclidean |
| **Naïve Bayes** | Distribution | Determines which distribution each feature is assumed to follow | Continuous features = Gaussian distribution. Categorical features= multinomial distribution |

[a] Specifies the parameter space for the random search strategy. Based on the results of the random search, a refined grid search across 500 steps was specified.

[b] Number of neurons in each hidden layer, where (1,) represents 1 neuron in the first hidden layer, with no further hidden layers; and (1,2) represents 1 neuron in the first hidden layer and 2 in the second hidden layer.

[c] The naïve Bayes algorithm did not undergo any hyperparameter grid search but instead required variables to be specified as either continuous and categorical at the time of model development.

Table E3 Comparability between predictors definitions in the IOWBC and MAAS cohorts

| Variable | IOWBC definition | MAAS definition | Comparability [a] |
|---|---|---|---|
| **Maternal age** | Maternal age at booking | Maternal age at birth of child | |
| **Birthweight** | Birth weight (kg) | Birthweight (kg) | |
| **Total breastfeeding** | Total breastfeeding duration | Breast feeding duration | |
| **Age of solid food introduction** | Age of introduction of cereals/solids (weeks) | At what age did your baby begin solid foods? (Weeks) | |
| **Early life BMI** | BMI at age 1, standardised against the British 1990 growth reference | SDS BMI at age 1, standardised against the British 1990 growth reference | |
| **Early life wheeze** | Frequency of asthma wheezing episodes at either 1 or 2 years | If no to: has or does your child's chest ever wheeze or whistle, what best describes your child's wheezing (at either 1 or 3 years) | **IOWBC:** categorised as no wheeze, occasional, frequent<br>**MAAS:** categorised as no wheeze, 1-2 times or from time to time (occasional), every day (frequent) |
| **Early life cough** | Asthmatic cough at either 1 or 2 years | Does your child usually have a cough apart from with colds at 1 or 3 years | |
| **Preschool BMI** | SDS BMI at age 4 | SDS BMI at age 5 | |
| **Preschool wheeze** | Frequency of wheezing at 4YR | Current wheeze age 5 years | |
| **Preschool cough** | Any asthmatic cough at 4 YR | Does your child usually have a cough during the day apart from with colds? | |
| **Preschool nocturnal symptoms** | Any nocturnal symptoms at 4YR | Does your child usually have a cough at night apart from with colds? Or, in the last 12 months how often - on average - has your child's sleep been disturbed by wheezing | |

| Variable | IOWBC definition | MAAS definition | Comparability [a] |
|---|---|---|---|
| **Preschool atopy** | Sensitisation (+SPT) to one or more allergens at age 4 | Sensitisation (+SPT) to one or more allergens at age 5 | **IOWBC**: tested allergens included house dust mite, milk, egg, cat, dog, grass, wheat, soya, peanut, cod, Cladosporium, Alternaria<br>**MAAS:** tested allergens included house dust mite, cat, dog, pollen, mould, milk, egg |
| **Preschool polysensitisation** | Sensitisation (+SPT) to two or more allergens by age 4 | Sensitisation (+SPT) to two or more allergens by age 5 | **IOWBC**: tested allergens included house dust mite, milk, egg, cat, dog, grass, wheat, soya, peanut, cod, Cladosporium, Alternaria<br>**MAAS:** tested allergens included house dust mite, cat, dog, pollen, mould, milk, egg |
| **Maternal socioeconomic status** | Maternal socioeconomic status | Maternal socioeconomic status | **IOWBC**: categorised into the following income strata: very low, low, low-middle, middle and high.<br>**MAAS**: categorised as routine (low), intermediate (low-middle), managerial (middle) and professional (high). |
| **School-age asthma** | Doctor diagnose asthma PLUS wheeze in the last 12 month AND/OR asthma treatment | Doctor diagnose asthma PLUS wheeze in the last 12 month AND/OR asthma treatment | **IOWBC:** evaluated at age 10<br>**MAAS:** evaluated at ages 8 and 11 |

[a] MAAS variable categorisations are given as: original categorisation of the MAAS variables (IOWBC equivalent used in the validation analysis).

Table E4 Descriptive statistics for all candidate features in the IOWBC analysed and the subset of individuals with complete data used for feature selection for the CAPE and CAPP prediction models

| | | Total IOWBC (n=1368) | | CAPE complete dataset (n=490) | | CAPP complete dataset (n=373) | |
|---|---|---|---|---|---|---|---|
| | | Asthmatic (n=201) | Non-asthmatic (n=1167) | Asthmatic (n=70) | Non-asthmatic (n=420) | Asthmatic (n=55) | Non-asthmatic (n=318) |
| **Family history** | **Maternal smoking at birth** | | | | | | |
| | No | 152 (75.62) | 877 (75.15) | 55 (78.57) | 345 (82.14) | 43 (78.18) | 257 (80.82) |
| | Yes | 47 (23.38) | 276 (23.65) | 15 (21.43) | 75 (17.86) | 12 (21.82) | 61 (19.18) |
| | **Paternal smoking at birth** | | | | | | |
| | No | 119 (59.20) | 714 (61.18) | 46 (65.71) | 283 (67.39) | 34 (61.82) | 206 (64. |
| | Yes | 79 (39.30) | 440 (37.70) | 24 (34.29) | 137 (32.62) | 21 (38.18) | 112 (35.22) |
| | **Maternal asthma** | | | | | | |
| | No | 170 (84.58) | 1047 (89.72) | 65 (92.86) | 381 (90.71) | 52 (94.55) | 293 (92.14) |
| | Yes | 29 (14.43) | 113 (9.68) | 5 (7.14) | 39 (9.29) | 3 (5.45) | 25 (7.86) |
| | **Maternal eczema** | | | | | | |
| | No | 170 (84.58) | 1025 (87.83) | 59 (84.29) | 371 (88.33) | 45 (81.82) | 284 (89.31) |
| | Yes | 28 (13.93) | 133 (11.40) | 11 (15.71) | 49 (11.67) | 10 (18.18) | 34 (10.69) |
| | **Maternal hay fever** | | | | | | |
| | No | 149 (74.13) | 941 (80.63) | 57 (81.43) | 335 (79.76) | 43 (78.18) | 256 (80.5) |
| | Yes | 50 (24.88) | 219 (18.77) | 13 (18.57) | 85 (20.24) | 12 (21.82) | 62 (19.50) |
| | **Paternal asthma** | | | | | | |
| | No | 171 (85.07) | 1049 (89.89) | 60 (85.71) | 387 (92.14) | 47 (85.45) | 291 (91.51) |
| | Yes | 27 (13.43) | 104 (8.91) | 10 (14.29) | 33 (7.86) | 8 (14.55) | 27 (8.49) |
| | **Paternal eczema** | | | | | | |
| | No | 179 (89.05) | 1082 (92.72) | 60 (85.71) | 395 (94.05) | 48 (87.27) | 298 (93.71) |
| | Yes | 19 (9.45) | 70 (6.00) | 10 (14.29) | 25 (5.95) | 7 (12.73)* | 20 (6.29)* |
| | **Paternal hay fever** | | | | | | |
| | No | 163 (81.09) | 987 (84.58) | 56 (80.00) | 366 (87.14) | 42 (76.36) | 278 (87.42) |
| | Yes | 35 (17.41) | 166 (14.22) | 14 (20.00) | 54 (12.86) | 13 (23.64) | 40 (12.58) |

| | | Total IOWBC (n=1368) | | CAPE complete dataset (n=490) | | CAPP complete dataset (n=373) | |
|---|---|---|---|---|---|---|---|
| | | **Asthmatic (n=201)** | **Non-asthmatic (n=1167)** | **Asthmatic (n=70)** | **Non-asthmatic (n=420)** | **Asthmatic (n=55)** | **Non-asthmatic (n=318)** |
| | **Parity** | | | | | | |
| | No | 78 (38.81) | 415 (35.56) | 28 (40.00) | 180 (42.86) | 19 (34.55) | 138 (43.4) |
| | Yes | 95 (47.26) | 573 (49.10) | 42 (60.00) | 240 (57.14) | 36 (65.45) | 180 (56.60) |
| | **Maternal socioeconomic status** | | | | | | |
| | Very low | 25 (12.44) | 163 (13.97) | 11 (15.71) | 43 (10.24) | 10 (18.18) | 30 (9.43) |
| | Low | 35 (17.41) | 199 (17.05) | 12 (17.14) | 81 (19.29) | 8 (14.55) | 63 (19.81) |
| | Low-Mid | 62 (30.85) | 334 (28.62) | 18 (25.71) | 128 (30.48) | 14 (25.45) | 103 (32.39) |
| | Mid | 52 (26.37) | 320 (27.42) | 21 (30.00) | 135 (32.14) | 16 (29.09) | 99 (31.13) |
| | High | 13 (6.47) | 96 (8.23) | 8 (11.43) | 33 (7.86) | 7 (12.73) | 23 (7.23) |
| **Prenatal/at birth** | **Maternal age** | 201 (26.61, 5.44) | 1167 (27.04, 5.26) | 70 (27.44, 5.32) | 420 (27.60, 4.91) | 55 (27.98, 5.37) | 318 (27.69, 4.95) |
| | **Prematurity** | | | | | | |
| | Pre-term | 9 (4.48) | 32 (2.74) | 1 (1.43) | 7 (1.67) | 1 (1.82) | 4 (1.26) |
| | Term | 184 (91.54) | 1103 (94.52) | 67 (95.71) | 411 (97.86) | 53 (96.36) | 312 (98.11) |
| | Post-term | 3 (1.49) | 12 (1.03) | 2 (2.86) | 2 (0.48) | 1 (1.82) | 2 (0.63) |
| | **Caesarean delivery** | | | | | | |
| | No | 150 (74.63) | 857 (73.44) | 62 (88.57) | 381 (90.71) | 48 (87.27) | 287 (90.25) |
| | Yes | 18 (8.96) | 86 (7.37) | 8 (11.43) | 39 (9.29) | 7 (12.73) | 31 (9.75) |
| | **Total breastfeeding** | | | | | | |
| | Never | 46 (22.89) | 267 (22.88) | 18 (25.71) | 95 (22.62) | 15 (27.27) | 73 (22.96) |
| | <3months | 66 (32.84) | 352 (30.16) | 29 (41.43) | 137 (32.62) | 22 (40.00) | 105 (33.02) |
| | 3-6 months | 22 (10.95) | 164 (14.05) | 4 (5.71) | 69 (16.43) | 3 (5.45) | 56 (17.61) |
| | >6 months | 37 (18.41) | 264 (22.62) | 19 (27.14) | 119 (28.33) | 15 (27.27) | 84 (26.42) |
| | **Exclusive breastfeeding** | | | | | | |
| | Never | 55 (27.36) | 334 (28.62) | 25 (35.71) | 125 (29.76) | 21 (38.18) | 91 (28.62) |
| | <3 months | 85 (42.29) | 489 (41.90) | 35 (50.00) | 191 (45.48) | 27 (49.09) | 146 (45.91) |
| | >3 months | 31 (15.42) | 224 (19.19) | 10 (14.29) | 104 (24.76) | 7 (12.72) | 81 (25.47) |

| | | Total IOWBC (n=1368) | | CAPE complete dataset (n=490) | | CAPP complete dataset (n=373) | |
|---|---|---|---|---|---|---|---|
| | | **Asthmatic (n=201)** | **Non-asthmatic (n=1167)** | **Asthmatic (n=70)** | **Non-asthmatic (n=420)** | **Asthmatic (n=55)** | **Non-asthmatic (n=318)** |
| | **Age of solid food introduction in weeks** | 168 (14.36, 4.51) | 1026 (14.34, 4.12) | 70 (13.96, 4.08) | 420 (14.45, 4.08) | 55 (13.96, 4.24) | 318 (14.59, 4.07) |
| | **Birthweight** | 199 (3.34, 0.52)* | 1142 (3.44, 0.50)* | 70 (3.45, 0.51) | 420 (3.47, 0.52) | 55 (3.48, 0.56) | 318 (3.45, 0.49) |
| | **Sex** | * | * | | | | |
| | Male | 118 (58.71) | 578 (49.53) | 40 (57.14) | 191 (45.48) | 31 (56.36) | 143 (44.97) |
| | Female | 83 (41.29) | 589 (50.47) | 30 (42.86) | 229 (54.52) | 24 (43.64) | 175 (55.03) |
| | **Season of birth** | | | | | | |
| | Autumn | 38 (18.91) | 243 (20.82) | 13 (18.57) | 101 (24.05) | 12 (21.82) | 86 (27.04) |
| | Winter | 64 (31.84) | 382 (32.73) | 19 (27.14) | 117 (27.86) | 14 (25.45) | 77 (24.21) |
| | Spring | 51 (25.37) | 274 (23.48) | 19 (27.14) | 100 (23.81) | 15 (27.27) | 80 (25.16) |
| | Summer | 48 (23.88) | 268 (22.96) | 19 (27.14) | 102 (24.29) | 14 (25.45) | 75 (23.58) |
| | **Dog** | | | | | | |
| | No | 149 (74.13) | 815 (69.84) | 55 (78.57) | 301 (71.67) | 41 (74.55) | 226 (71.07) |
| | Yes | 51 (25.37) | 346 (29.65) | 15 (21.43) | 119 (28.33) | 14 (25.45) | 92 (28.93) |
| | **Cat** | | | | | | |
| | No | 143 (71.14) | 764 (65.47) | 46 (65.71) | 274 (65.24) | 35 (63.64) | 205 (64.47) |
| | Yes | 57 (28.36) | 397 (34.02) | 24 (34.29) | 146 (34.76) | 20 (36.36) | 113 (35.53) |
| | **Furry pet** | | | | | | |
| | No | 105 (52.24) | 525 (44.99) | 39 (55.71) | 194 (46.19) | 29 (52.73) | 143 (44.97) |
| | Yes | 95 (47.26) | 636 (54.50) | 31 (44.29) | 226 (53.81) | 26 (47.27) | 175 (55.03) |
| **Early life** | **BMI** | 135 (-0.15, 1.15) | 851 (-0.16, 1.22) | 70 (-0.13, 1.12) | 420 (-0.17, 1.19) | 55 (-0.20, 1.13) | 318 (-0.18, 1.19) |
| | **Wheeze** | * | * | * | * | * | * |
| | Never | 78 (38.81) | 739 (63.32) | 39 (55.71) | 341 (81.19) | 30 (54.55) | 259 (81.45) |
| | Occasional | 14 (6.97) | 63 (5.40) | 6 (8.57) | 28 (6.67) | 5 (9.09) | 20 (6.29) |
| | Frequent | 56 (27.86) | 124 (10.63) | 25 (35.71) | 51 (12.14) | 20 (36.36) | 39 (12.26) |

| | Total IOWBC (n=1368) | | CAPE complete dataset (n=490) | | CAPP complete dataset (n=373) | |
|---|---|---|---|---|---|---|
| | **Asthmatic (n=201)** | **Non-asthmatic (n=1167)** | **Asthmatic (n=70)** | **Non-asthmatic (n=420)** | **Asthmatic (n=55)** | **Non-asthmatic (n=318)** |
| **Wheeze without cold** | * | * | * | * | * | * |
| No | 89 (44.28) | 796 (68.21) | 45 (64.29) | 369 (87.86) | 35 (63.64) | 279 (87.74) |
| Yes | 56 (27.86) | 124 (10.63) | 25 (35.71) | 51 (12.14) | 20 (36.36) | 39 (12.26) |
| **Cough** | * | * | * | * | * | * |
| No | 78 (38.81) | 749 (64.18) | 39 (55.71) | 346 (82.38) | 30 (54.55) | 263 (82.7) |
| Yes | 70 (34.83) | 174 (14.91) | 31 (44.29) | 74 (17.62) | 25 (45.45) | 55 (17.30) |
| **Nasal symptoms** | * | * | * | * | * | * |
| No | 101 (50.25) | 757 (64.87) | 45 (64.29) | 331 (78.81) | 36 (65.45) | 252 (79.25) |
| Yes | 60 (29.85) | 239 (20.48) | 25 (35.71) | 89 (21.19) | 19 (34.55) | 66 (20.75) |
| **Chest infection** | * | * | * | * | * | * |
| No | 103 (51.24) | 830 (71.12) | 51 (72.86) | 375 (89.29) | 40 (72.73) | 284 (89.31) |
| Yes | 54 (26.87) | 139 (11.91) | 19 (27.14) | 45 (10.71) | 15 (27.27) | 34 (10.69) |
| **Nocturnal symptoms** | * | * | * | * | * | * |
| No | 80 (39.80) | 756 (64.78) | 41 (58.57) | 347 (82.62) | 32 (58.18) | 264 (83.02) |
| Yes | 68 (33.83) | 161 (13.80) | 29 (41.43) | 73 (17.28) | 23 (41.82) | 54 (16.98) |
| **Eczema** | * | * | * | * | | |
| No | 95 (47.26) | 740 (63.41) | 46 (65.71) | 320 (76.19) | 35 (63.64) | 246 (77.36) |
| Yes | 66 (32.84) | 247 (21.17) | 24 (34.29) | 100 (23.81) | 20 (36.36) | 72 (22.64) |
| **Hay fever** | | | | | | |
| No | 123 (61.19) | 827 (70.87) | 53 (75.71) | 353 (84.05) | 42 (76.36) | 268 (84.28) |
| Yes | 36 (17.91) | 165 (14.14) | 17 (24.29) | 67 (15.95) | 13 (23.64) | 50 (15.72) |
| **Atopy** | * | * | * | * | * | * |
| No | 95 (47.26) | 834 (71.47) | 53 (75.71) | 392 (93.33) | 43 (78.18) | 304 (95.6) |
| Yes | 44 (21.89) | 58 (4.97) | 17 (24.29) | 28 (6.67) | 12 (21.82) | 14 (4.40) |
| **Monosensitisation** | * | * | * | * | * | * |
| No | 99 (49.25) | 840 (71.98) | 55 (78.57) | 394 (93.81) | 44 (80) | 305 (95.91) |
| Yes | 37 (18.41) | 51 (4.37) | 15 (21.43) | 26 (6.19) | 11 (20.00) | 13 (4.09) |

| | | | Total IOWBC (n=1368) | | CAPE complete dataset (n=490) | | CAPP complete dataset (n=373) | |
|---|---|---|---|---|---|---|---|---|
| | | | **Asthmatic (n=201)** | **Non-asthmatic (n=1167)** | **Asthmatic (n=70)** | **Non-asthmatic (n=420)** | **Asthmatic (n=55)** | **Non-asthmatic (n=318)** |
| | **Polysensitisation** | | * | * | * | * | * | * |
| | | No | 119 (59.20) | 876 (75.06) | 67 (95.71) | 417 (99.29) | 53 (96.36) | 317 (99.69) |
| | | Yes | 10 (4.98) | 9 (0.77) | 3 (4.29) | 3 (0.71) | 2 (3.64) | 1 (0.31) |
| | **Parental smoking** | | | | | | | |
| | | Never | 69 (34.33) | 470 (40.27) | 33 (47.14) | 231 (55.00) | 24 (43.64) | 168 (52.83) |
| | | Ex-smoker | 5 (2.49) | 54 (4.63) | 2 (2.86) | 29 (6.90) | 2 (3.64) | 23 (7.23) |
| | | Current | 93 (46.27) | 488 (41.82) | 35 (50.00) | 160 (38.10) | 29 (52.73) | 127 (39.94) |
| | **Dog** | | | | | | | |
| | | No | 118 (58.71) | 656 (56.21) | 55 (78.57) | 294 (70.00) | 41 (74.55) | 219 (68.87) |
| | | Yes | 41 (20.40) | 327 (28.02) | 15 (21.43) | 126 (30.00) | 14 (25.45) | 99 (31.13) |
| | **Cat** | | | | | | | |
| | | No | 26 (12.94) | 176 (15.08) | 11 (15.71) | 71 (16.90) | 9 (16.36) | 54 (16.98) |
| | | Yes | 131 (65.17) | 821 (70.35) | 59 (84.29) | 349 (83.10) | 46 (83.64) | 264 (83.02) |
| | **Furry pet** | | | | | | | |
| | | No | 6 (2.99) | 19 (1.63) | 4 (5.71) | 8 (1.90) | 3 (5.45) | 7 (2.20) |
| | | Yes | 155 (77.11) | 1001 (85.78) | 66 (94.29) | 412 (98.10) | 52 (94.55) | 311 (97.80) |
| | **Early life residence on a farm** | | | | | | | |
| | | No | 174 (86.57) | 990 (84.83) | 66 (94.29) | 398 (94.76) | 52 (94.55) | 305 (95.91) |
| | | Yes | 6 (2.99) | 43 (3.68) | 4 (5.71) | 22 (5.24) | 3 (5.45) | 13 (4.09) |
| **Preschool age** | **BMI** | | 146 (0.21, 1.03) | 855 (0.23, 1.04) | - | - | 55 (0.28, 0.88) | 318 (0.28, 0.95) |
| | **Wheeze** | | * | * | | | * | * |
| | | Never | 85 (42.29) | 879 (75.32) | - | - | 28 (50.91) | 281 (88.36) |
| | | Occasional | 18 (8.96) | 34 (2.91) | - | - | 7 (12.73) | 10 (3.14) |
| | | Frequent | 70 (34.83) | 75 (6.43) | - | - | 20 (36.36) | 27 (8.49) |
| | **Wheeze without cold** | | * | * | | | * | * |
| | | No | 103 (51.24) | 913 (78.23) | - | - | 35 (63.64) | 291 (91.51) |
| | | Yes | 70 (34.83) | 75 (6.43) | - | - | 20 (36.36) | 27 (8.49) |

| | Total IOWBC (n=1368) | | CAPE complete dataset (n=490) | | CAPP complete dataset (n=373) | |
|---|---|---|---|---|---|---|
| | Asthmatic (n=201) | Non-asthmatic (n=1167) | Asthmatic (n=70) | Non-asthmatic (n=420) | Asthmatic (n=55) | Non-asthmatic (n=318) |
| **Cough** | * | * | | | * | * |
| No | 74 (36.82) | 860 (73.69) | - | - | 23 (41.82) | 279 (87.74) |
| Yes | 99 (49.25) | 128 (10.97) | - | - | 32 (58.18)* | 39 (12.26)* |
| **Nasal symptoms** | * | * | | | * | * |
| No | 112 (55.72) | 852 (73.01) | - | - | 34 (61.82) | 275 (86.48) |
| Yes | 61 (30.35) | 136 (11.65) | - | - | 21 (38.18)* | 43 (13.52)* |
| **Nocturnal symptoms** | * | * | | | * | * |
| No | 79 (39.30) | 860 (73.69) | - | - | 25 (45.45) | 277 (87.11) |
| Yes | 94 (46.77) | 128 (10.97) | - | - | 30 (54.55)* | 41 (12.89)* |
| **Eczema** | * | * | | | | |
| No | 126 (62.69) | 901 (77.21) | - | - | 45 (81.82) | 290 (91.19) |
| Yes | 47 (23.38) | 87 (7.46) | - | - | 10 (18.18) | 28 (8.81) |
| **Hay fever** | * | * | | | * | * |
| No | 144 (71.64) | 953 (81.66) | - | - | 45 (81.82) | 303 (95.28) |
| Yes | 29 (14.43) | 35 (3.00) | - | - | 10 (18.18)* | 15 (4.72)* |
| **Atopy** | * | * | | | * | * |
| No | 67 (33.33) | 670 (57.41) | - | - | 29 (52.73) | 275 (86.48) |
| Yes | 72 (35.82) | 94 (8.05) | - | - | 26 (47.27)* | 43 (13.52)* |
| **Monosensitisation** | * | * | | | | |
| No | 115 (57.21) | 708 (60.67) | - | - | 46 (83.64) | 292 (91.82) |
| Yes | 22 (10.95) | 53 (4.54) | - | - | 9 (16.36) | 26 (8.18) |
| **Polysensitisation** | * | * | | | * | * |
| No | 93 (46.27) | 874 (74.89) | - | - | 38 (69.09) | 301 (94.65) |
| Yes | 48 (23.88) | 38 (3.26) | - | - | 17 (30.91)* | 17 (5.35)* |

| | Total IOWBC (n=1368) | | CAPE complete dataset (n=490) | | CAPP complete dataset (n=373) | |
|---|---|---|---|---|---|---|
| | **Asthmatic (n=201)** | **Non-asthmatic (n=1167)** | **Asthmatic (n=70)** | **Non-asthmatic (n=420)** | **Asthmatic (n=55)** | **Non-asthmatic (n=318)** |
| **Parental smoking** | | | | | | |
| Never | 62 (30.85) | 424 (36.33) | - | - | 22 (40.00) | 157 (49.37) |
| Ex-smoker | 19 (9.45) | 122 (10.45) | - | - | 6 (10.91) | 47 (14.78) |
| Current | 78 (38.81) | 357 (30.59) | - | - | 27 (49.09) | 114 (35.85) |
| **Dog** | | | | | | |
| No | 125 (62.19) | 710 (60.84) | - | - | 40 (72.73) | 227 (71.38) |
| Yes | 50 (24.88) | 280 (23.99) | - | - | 15 (27.27) | 91 (28.62) |
| **Cat** | | | | | | |
| No | 115 (57.21) | 620 (53.13) | - | - | 40 (72.73) | 198 (62.26) |
| Yes | 60 (29.85) | 370 (31.71) | - | - | 15 (47.27) | 120 (37.74) |
| **Furry pet** | | | | | | |
| No | 75 (37.31) | 386 (33.08) | - | - | 18 (32.73) | 123 (38.68) |
| Yes | 96 (47.76) | 586 (50.21) | - | - | 37 (67.27) | 195 (61.32) |

Summary data is reported as the number of individuals with data, with the mean and standard deviation ($\bar{x}$, s) for the continuous features of: maternal age, birthweight, age of solid food introduction, early life BMI and preschool BMI; or proportions for the remaining categorical features (%). Where the number of individuals with data for a variable does not equal the total number of individuals detailed in the column, the difference indicates the number of individuals with missing data.

* Indicates a statistically significant difference for the variable between asthmatic and non-asthmatic children at age 10 ($p<0.05$), assessed using an independent two sample t-test or Pearson's Chi-square test for independence for continuous and categorical features, respectively.

Table E5 Directionality of school-age asthma risk incurred by each selected model predictor

| Predictor | Log odds (95% CI) | P-value | Effect on asthma risk |
|---|---|---|---|
| **Maternal age** | -0.02 (-0.04, 0.01) | 0.28 | Negligible [a] |
| **Birthweight** | -0.38 (-0.68, -0.09) | 0.01 | Decrease [a] |
| **Age of solid food introduction** | 0.00 (-0.04, 0.04) | 0.96 | Negligible [a] |
| **Breastfeeding duration** | -0.09 (-0.24, 0.05) | 0.22 | Negligible [a] |
| **Early life BMI** | 0.00 (-0.14, 0.16) | 0.92 | Negligible [a] |
| **Preschool BMI** | -0.01 (-0.18, 0.16) | 0.88 | Negligible [a] |
| **Early life wheeze** | 0.73 (0.53, 0.92) | <0.01 | Increase |
| **Early life cough** | 1.35 (0.99, 1.71) | <0.01 | Increase |
| **Preschool wheeze** | 1.15 (0.96, 1.35) | <0.01 | Increase |
| **Preschool cough** | 2.20 (1.84, 2.55) | <0.01 | Increase |
| **Preschool nocturnal symptoms** | 2.08 (1.73, 2.43) | <0.01 | Increase |
| **Preschool atopy status** | 2.04 (1.64, 2,44) | <0.01 | Increase |
| **Preschool polysensitisation status** | 2.47 ( 2.00, 2.96) | <0.01 | Increase |
| **Maternal socioeconomic status** | -0.01 (-0.15, 0.12) | 0.83 | Negligible [b] |

Univariate logistic regression of the effect of each feature selected by RFE (for use in either the infancy and preschool models) on school-age asthma development at age 10.

95% CI = 95% confidence interval

[a] The effect on asthma risk is evaluated based on the increasing value of the predictor e.g. higher birthweight is suggested to decrease the asthma risk.

[b] The effect on asthma risk is based on an increase in maternal socioeconomic status

Table E6 Distribution of CAPE and CAPP model predictors for individuals in the IOWBC and MAAS at each asthma prediction timepoint.

| | IOWBC 10YR (n=1368) | | MAAS 8YR (n=1018) | | MAAS 11YR (n=898) | |
|---|---|---|---|---|---|---|
| | **Asthmatic (n=201)** | **Non-asthmatic (n=1167)** | **Asthmatic (n=144)** | **Non-asthmatic (n=874)** | **Asthmatic (n=116)** | **Non-asthmatic (n=782)** |
| **Maternal age** | 201 (26.61, 5.44) | 1167 (27.04, 5.26) | 116 (30.53, 5.09) | 842 (20.66, 4.67) | 94 (29.59, 4.94))* | 762 (30.88, 4.61)* |
| **Birthweight** | 199 (3.34, 0.52)* | 1142 (3.44, 0.50)* | 132 (3.44, 0.50) | 845 (3.49, 0.49) | 107 (3.41, 0.51) | 757 (3.49, 0.49) |
| **Age of solid food introduction** | 168 (14.36, 4.51) | 1026 (14.34, 4.12) | 51 (14.88, 3.83) | 392 (14.67, 3.52) | 44 (14.93, 5.03) | 351 (14.69, 3.34) |
| **Breastfeeding duration** | | | | | | |
| Never | 46 (22.89) | 267 (22.88) | 47 (23.64) | 281 (32.15) | 32 (27.59) | 236 (30.18) |
| <3months | 66 (32.84) | 352 (30.16) | 33 (22.92) | 214 (24.49) | 30 (25.86) | 190 (24.30) |
| 3-6 months | 22 (10.95) | 164 (14.05) | 24 (16.67) | 162 (18.54) | 16 (12.79) | 157 (20.08) |
| >6 months | 37 (18.41) | 264 (22.62) | 22 (15.28) | 194 (22.20) | 23 (19.83) | 181 (23.15) |
| **Early life BMI** | 135 (-0.15, 1.15) | 851 (-0.16, 1.22) | 49 (-0.18, 1.00) | 387 (-0.25, 1.11) | 43 (-0.04, 1.09) | 347 (-0.32, 1.12) |
| **Preschool BMI** | 146 (0.21, 1.03) | 855 (0.23, 1.04) | 134 (0.57, 0.95) | 804 (0.46, 0.94) | 113 (0.65, 0.90)* | 731 (0.42, 0.94)* |
| **Early life wheeze** | * | * | * | * | * | * |
| Never | 78 (38.81) | 739 (63.32) | 8 (5.56) | 212 (24.26) | 9 (7.76) | 189 (24.17) |
| Occasional | 14 (6.97) | 63 (5.40) | 93 (64.58) | 339 (38.79) | 67 (57.76) | 299 (38.24) |
| Frequent | 56 (27.86) | 124 (10.63) | 4 (2.78) | 6 (0.69) | 4 (3.45) | 5 (0.64) |
| **Early life cough** | * | * | * | * | * | * |
| No | 78 (38.81) | 749 (64.18) | 31 (21.53) | 318 (36.38) | 28 (24.14) | 281 (35.93) |
| Yes | 70 (34.83) | 174 (14.91) | 34 (23.61) | 135 (15.45) | 32 (27.59) * | 117 (14.96) * |
| **Preschool wheeze** | * | * | * | * | * | * |
| Never | 85 (42.29) | 879 (75.32) | 44 (30.56) | 721 (82.49) | 40 (34.48) | 655 (83.76) |
| Occasional | 18 (8.96) | 34 (2.91) | 89 (61.81) | 116 (13.27) | 66 (56.90) | 105 (13.43) |
| Frequent | 70 (34.83) | 75 (6.43) | 5 (3.47) | 6 (0.69) | 6 (5.17) | 3 (0.38) |
| **Preschool cough** | * | * | * | * | * | * |
| No | 74 (36.82) | 860 (73.69) | 70 (48.61) | 711 (81.35) | 62 (53.45) | 642 (82.10) |
| Yes | 99 (49.25) | 128 (10.97) | 68 (47.22) | 132 (15.10) | 50 (43.10) | 121 (15.47) |

| | IOWBC 10YR (n=1368) | | MAAS 8YR (n=1018) | | MAAS 11YR (n=898) | |
|---|---|---|---|---|---|---|
| | Asthmatic (n=201) | Non-asthmatic (n=1167) | Asthmatic (n=144) | Non-asthmatic (n=874) | Asthmatic (n=116) | Non-asthmatic (n=782) |
| **Preschool nocturnal symptoms** | * | * | * | * | * | * |
| No | 79 (39.30) | 860 (73.69) | 49 (34.03) | 700 (80.09) | 47 (40.52) | 629 (80.43) |
| Yes | 94 (46.77) | 128 (10.97) | 89 (61.81) | 143 (16.36) | 65 (56.03) | 134 (17.14) |
| **Preschool atopy status** | * | * | * | * | * | * |
| No | 67 (33.33) | 670 (57.41) | 52 (36.11) | 573 (65.56) | 38 (32.76) | 530 (67.77) |
| Yes | 72 (35.82) | 94 (8.05) | 76 (52.78) | 197 (22.54) | 69 (59.48) | 169 (21.61) |
| **Preschool polysensitisation status** | * | * | * | * | * | * |
| No | 93 (46.27) | 874 (74.89) | 77 (53.47) | 675 (77.23) | 62 (53.45) | 612 (78.26) |
| Yes | 48 (23.88) | 38 (3.26) | 47 (32.64) | 77 (8.81) | 41 (35.34) | 73 (9.34) |
| **Maternal socioeconomic status** | | | | | | |
| Very low | 25 (12.44) | 163 (13.97) | 0 (0.00) | 0 (0.00) | 0 (0.00) | 0 (0.00) |
| Low | 35 (17.41) | 199 (17.05) | 11 (7.64) | 66 (7.55) | 9 (7.76) | 54 (6.91) |
| Low-Mid | 62 (30.85) | 334 (28.62) | 15 (10.42) | 137 (15.68) | 10 (8.62) | 129 (16.50) |
| Mid | 52 (26.37) | 320 (27.42) | 41 (28.47) | 388 (44.39) | 30 (25.86) | 375 (47.95) |
| High | 13 (6.47) | 96 (8.23) | 0 (0.00) | 0 (0.00) | 0 (0.00) | 0 (0.00) |

Summary data for predictors included in the CAPE and CAPP models is reported for all individuals with a reported asthma status in the IOWBC or MAAS at each prediction time point.

The distribution of predictors is reported as the number of individuals, with mean and standard deviation ($\bar{x}$, s) for the continuous features of: maternal age, birthweight, age of solid food introduction, early life BMI and preschool BMI; or as proportions for the remaining categorical features (%). Where the number of individuals with data for a variable does not equal the total number of individuals detailed in the column, the difference indicates the number of individuals with missing data.

* Statistically significant difference between asthmatic and non-asthmatic children ($p<0.05$), assessed using an independent two sample t-test or Pearson's Chi-square test for independence for continuous and categorical features, respectively.

Table E7 Model performance for predicting an alternative definition of asthma

| | Asthma definition[a] (% asthmatic) | Balanced accuracy | AUC | Sensitivity | Specificity | PPV | NPV | LR+ | LR- | F1 score |
|---|---|---|---|---|---|---|---|---|---|---|
| CAPE Model | IOWBC Original (13.3%) | 0.71 (0.62-0.78) | 0.71 (0.61-0.80) | 0.74 (0.56-0.88) | 0.68 (0.62-0.74) | 0.26 (0.21-0.32) | 0.94 (0.91-0.97) | 2.29 (1.69-3.01) | 0.39 (0.18-0.63) | 0.38 (0.31-0.46) |
| | IOWBC Alternative (8.1%) | 0.70 (0.62-0.76) | 0.67 (0.56-0.78) | 0.90 (0.75-1.00) | 0.49 (0.43-0.56) | 0.13 (0.11-0.16) | 0.98 (0.96-1.00) | 1.77 (1.44-2.12) | 0.20 (0.00-0.49) | 0.23 (0.20-0.27) |
| | MAAS 8YR Alternative (5.0%) | 0.61 (0.51-0.68) | 0.69 (0.55-0.82) | 0.87 (0.67-1.00) | 0.34 (0.29-0.40) | 0.07 (0.05-0.08) | 0.98 (0.95-1.00) | 1.32 (1.02-1.57) | 0.39 (0.00-0.97) | 0.12 (0.09-0.14) |
| | MAAS 11YR Alternative (3.0%) | 0.60 (0.45-0.69) | 0.58 (0.37-0.75) | 0.86 (0.57-1.00) | 0.33 (0.28-0.39) | 0.03 (0.02-0.04) | 0.99 (0.97-1.00) | 1.29 (0.84-1.61) | 0.43 (0.00-1.34) | 0.06 (0.04-0.08) |
| CAPP Model | IOWBC Original (13.7%) | 0.80 (0.70-0.89) | 0.82 (0.71-0.91) | 0.72 (0.52-0.88) | 0.88 (0.83-0.92) | 0.47 (0.38-0.62) | 0.95 (0.92-0.98) | 5.99 (3.79-10.11) | 0.32 (0.13-0.54) | 0.56 (0.45-0.70) |
| | IOWBC Alternative (10.1%) | 0.77 (0.68-0.85) | 0.79 (0.67-0.89) | 0.83 (0.67-1.00) | 0.71 (0.64-0.78) | 0.25 (0.19-0.31) | 0.97 (0.95-1.00) | 2.92 (2.11-4.07) | 0.23 (0.00-0.48) | 0.38 (0.30-0.46) |
| | MAAS 8YR Alternative (5.3%) | 0.68 (0.56-0.78) | 0.70 (0.57-0.82) | 0.79 (0.57-1.00) | 0.57 (0.51-0.63) | 0.09 (0.07-0.12) | 0.98 (0.96-1.00) | 1.83 (1.29-2.39) | 0.38 (0.00-0.77) | 0.17 (0.12-0.21 |
| | MAAS 11YR Alternative (2.4%) | 0.71 (0.53-0.81) | 0.68 (0.40-0.87) | 0.83 (0.50-1.00) | 0.58 (0.52-0.64) | 0.05 (0.03-0.06) | 0.99 (0.98-1.00) | 1.98 (1.15-2.63) | 0.29 (0.00-0.88) | 0.09 0.05-0.12) |

[a] The outcome of school-age asthma was defined as follows: original asthma definition= doctor diagnosis of asthma ever plus the presence of wheeze or use of asthma medication in the last 12 months; alternative asthma definition= current wheeze and bronchial hyper-responsiveness. Both asthma outcomes were evaluated at age 10 in the IOWBC, in individuals in the respective test sets for each model.

[b] The final column presents the confusion matrix for the model classifications, where TN=true negatives, FP=false positives, FN=false negatives, TP=true positives.

Table E8 Reclassification table comparing predictions made by the PARS and CAPP models in MAAS

| | | Predicted risk (CAPP model) | | | Reclassified by CAPP (%) | | | |
|---|---|---|---|---|---|---|---|---|
| | **Predicted risk (PARS model)** | **No asthma** | **Asthma** | **Total** | **Increased risk** | **Decreased risk** | **Correctly reclassified** | **NRI** |
| **MAAS 8YR (PARS AUC=0.86 vs CAPP=0.83)** | | | | | | | | |
| **No asthma (n=213)** | **No asthma** | 173 | 14 | 187 | | | | |
| | **Asthma** | 21 | 5 | 26 | 14(7%) | 21(10%) | 21(10%) | 0.03 |
| | **Total** | 194 | 19 | 213 | | | | |
| **Asthma (n=28)** | **No asthma** | 5 | 7 | 12 | | | | |
| | **Asthma** | 7 | 9 | 16 | 7(25%) | 7(25%) | 7(25%) | 0.00 |
| | **Total** | 12 | 16 | 28 | 21 | 28 | 28 | |
| **MAAS 11YR (PARS AUC=0.78 vs CAPP=0.79)** | | | | | | | | |
| **No asthma (n=215)** | **No asthma** | 170 | 14 | 184 | | | | |
| | **Asthma** | 24 | 7 | 31 | 14(7%) | 24(11%) | 24(11%) | 0.05 |
| | **Total** | 194 | 21 | 215 | | | | |
| **Asthma (n=26)** | **No asthma** | 8 | 7 | 15 | | | | |
| | **Asthma** | 4 | 7 | 11 | 7(27%) | 4(15%) | 7(27%) | 0.12 |
| | **Total** | 12 | 14 | 26 | 21 | 28 | 21 | |

Table E9 Comparison of the performance of the CAPE and CAPP tools with their benchmark regression-based models

| Model | AUC | Sensitivity | Specificity | PPV | NPV | LR+ | LR- |
|---|---|---|---|---|---|---|---|
| **CAPE in IOW** | 0.71 | 0.74 | 0.68 | 0.26 | 0.94 | 2.29 | 0.39 |
| **CAPP in IOW** | 0.82 | 0.72 | 0.88 | 0.47 | 0.95 | 5.99 | 0.32 |
| **Loose API** | - | 0.42 | 0.85 | 0.59 | 0.73 | 2.72[a] | 0.69[a] |
| **Stringent API** | - | 0.16 | 0.97 | 0.76 | 0.68 | 6.04[a] | 0.87[a] |
| **PIAMA** | 0.74 | 0.19 | 0.97 | 0.42 | 0.91 | 6.33[a] | 0.84[a] |
| **PAPS** | 0.66 | 0.42 | 0.90 | 0.67 | 0.76 | 4.06 | 0.64 |
| **PARC** | 0.74 | 0.72 | 0.71 | 0.49 | 0.86 | 2.47 | 0.40 |
| **PARS** | 0.80 | 0.68 | 0.77 | 0.37 | 0.93 | 3.02 | 0.41 |

Predictive performance of the published validated studies compared to the machine learning CAPE and CAPP models developed in this study.

For each model, performance measures are reported based on the optimal threshold specified in their original papers.

[a]Performance measures were not reported in the original study so were calculated to enable comparison.

Table E10 Performance of the CAPP Tool with and without the predictors of sensitisation

| | Dataset | Balanced Accuracy | AUC | Sensitivity | Specificity | PPV | NPV | LR+ | LR- | F1 Score |
|---|---|---|---|---|---|---|---|---|---|---|
| **CAPP Model** | **IOWBC** | 0.80 (0.70-0.89) | 0.82 (0.71-0.91) | 0.72 (0.52-0.88) | 0.88 (0.83-0.92) | 0.47 (0.38-0.62) | 0.95 (0.92-0.98) | 5.99 (3.79-10.11) | 0.32 (0.13-0.54) | 0.56 (0.45-0.70) |
| | **MAAS 8YR** | 0.73 (0.64-0.81) | 0.83 (0.75-0.90) | 0.55 (0.36-0.70) | 0.91 (0.88-0.95) | 0.45 (0.33-0.59) | 0.94 (0.92-0.96) | 6.17 (3.64-10.69) | 0.50 (0.33-0.69) | 0.49 (0.36-0.62) |
| | **MAAS 11YR** | 0.73 (0.63-0.82) | 0.79 (0.68-0.88) | 0.55 (0.38-0.72) | 0.90 (0.87-0.94) | 0.41 (0.29-0.55) | 0.94 (0.92-0.96) | 5.71 (3.44-9.85) | 0.50 (0.30-0.71) | 0.47 (0.33-0.62) |
| **CAPP Model without sensitisation** | **IOWBC** | 0.75 (0.64-0.84) | 0.72 (0.58-0.85) | 0.64 (0.44-0.80) | 0.85 (0.80-0.91) | 0.41 (0.30-0.53) | 0.94 (0.91-0.97) | 4.40 (2.71-7.22) | 0.42 (0.22-0.66) | 0.50 (0.36-0.63) |
| | **MAAS 8YR** | 0.67 (0.59-0.76) | 0.79 (0.70-0.87) | 0.47 (0.31-0.64) | 0.87 (0.83-0.91) | 0.33 (0.23-0.44) | 0.92 (0.90-0.95) | 3.69 (2.23-5.91) | 0.61 (0.41-0.80) | 0.39 (0.27-0.51) |
| | **MAAS 11YR** | 0.65 (0.56-0.74) | 0.70 (0.57-0.81) | 0.42 (0.26-0.58) | 0.87 (0.83-0.91) | 0.29 (0.19-0.40) | 0.92 (0.90-0.95) | 3.32 (1.87-5.55) | 0.66 (0.46-0.87) | 0.34 (0.21-0.47) |

### Supplementary Figures

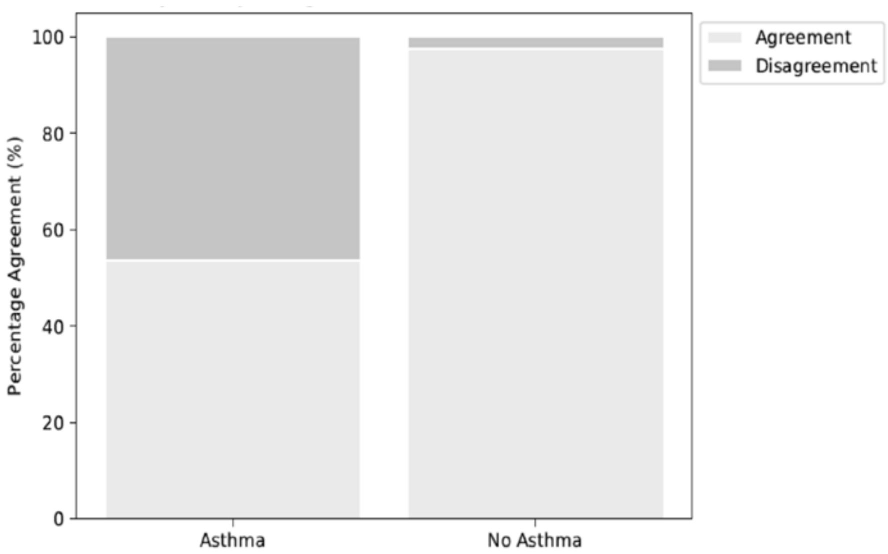

Figure E1 Agreement between the original and modified asthma definitions

Of the 1368 individuals in the IOWBC included in the main study, 1312 individuals had their asthma status defined using the two asthma definitions: original definition used in the analysis (doctor diagnosis asthma ever and wheeze or use of asthma medication in the last 12 months) and a modified definition (wheeze in the last 12 months and BHR). Each stacked bar represents the classification of individuals as asthmatic (left, n=160) or non-asthmatic (right, n=1152) based on the original asthma definition. Each bar shows the proportion of individuals for whom the modified asthma definition assigned the same asthma status (green stacks) or opposing asthma status (orange stacks) compared to the original asthma definition.

| **CAPE Tool Questionnaire (use if the child is an infant)** | |
|---|---|
| What was your child’s birthweight? | __ kg |
| At what age was your child introduced to solid foods? | __ weeks |
| Did you breastfeed your child? If yes, how long did you breastfeed for? | __ months |
| What was your child’s BMI when they were one year old? | __ |
| How often did your child experience any wheezing or whistling in their first two years of life? | ☐ Never<br>☐ 1-3 times per year<br>☐ 3+ times per year |
| Did your child experience coughing in the absence of a cold in their first two years of life? | Yes/ No |
| How old were you (the mother) at the time of your child’s birth? | __ years |
| How would you best describe your socioeconomic class? | ☐ Very low<br>☐ Low<br>☐ Lower-Middle<br>☐ Middle<br>☐ Upper |

Figure E2 Parental questionnaire for collecting data needed for the CAPE tool

| | |
|---|---|
| **CAPP Tool Questionnaire (use if the child is of preschool age, 3-5 years old)** | |
| What was your child's birthweight? | __ kg |
| At what age was your child introduced to solid foods? | __ weeks |
| Did you breastfeed your child? If yes, how long did you breastfeed for? | __ months |
| What was your child's approximate height and weight when they were one year old? | __cm<br>__kg |
| How old were you (the mother) at the time of your child's birth? | __ years |
| How would you best describe your socioeconomic class? | ☐ Very low<br>☐ Low<br>☐ Lower-Middle<br>☐ Middle<br>☐ Upper |
| **Current status:** | |
| What is your child's approximate height and weight? | __cm<br>__kg |
| How often does your child experience any wheezing or whistling? | ☐ Never<br>☐ 1-3 times per year<br>☐ 3+ times per year |
| Does your child experience coughing in the absence of a cold? | Yes/ No |
| Does your child experience asthma-like symptoms (wheeze/cough) at night night? | Yes/ No |
| If your child has undergone a recent skin prick test, how many food/aeroallergens were they sensitised (positive) to? | __<br>(number of allergens or NA if test not done) |

Figure E3 Parental questionnaire for collecting data needed for the CAPP tool

### TRIPOD Checklist: Prediction Model Development and Validation

| Section/Topic | Ite | | Checklist Item | Page |
|---|---|---|---|---|
| **Title and abstract** | | | | |
| Title | 1 | D;V | Identify the study as developing and/or validating a multivariable prediction model, the target population, and the outcome to be predicted. | 3 |
| Abstract | 2 | D;V | Provide a summary of objectives, study design, setting, participants, sample size, predictors, outcome, statistical analysis, results, and conclusions. | 3 |
| **Introduction** | | | | |
| Background and objectives | 3a | D;V | Explain the medical context (including whether diagnostic or prognostic) and rationale for developing or validating the multivariable prediction model, including references to existing models. | 6-7 |
| | 3b | D;V | Specify the objectives, including whether the study describes the development or validation of the model or both. | 7 |
| **Methods** | | | | |
| Source of data | 4a | D;V | Describe the study design or source of data (e.g., randomized trial, cohort, or registry data), separately for the development and validation data sets, if applicable. | 8 |
| | 4b | D;V | Specify the key study dates, including start of accrual; end of accrual; and, if applicable, end of follow-up. | SM-4 |
| Participants | 5a | D;V | Specify key elements of the study setting (e.g., primary care, secondary care, general population) including number and location of centres. | 7, SM-4 |
| | 5b | D;V | Describe eligibility criteria for participants. | 8 |
| | 5c | D;V | Give details of treatments received, if relevant. | NA |
| Outcome | 6a | D;V | Clearly define the outcome that is predicted by the prediction model, including how and when assessed. | 8 |
| | 6b | D;V | Report any actions to blind assessment of the outcome to be predicted. | NA |
| Predictors | 7a | D;V | Clearly define all predictors used in developing or validating the multivariable prediction model, including how and when they were measured. | 8, SM-14 |
| | 7b | D;V | Report any actions to blind assessment of predictors for the outcome and other predictors. | 8 |
| Sample size | 8 | D;V | Explain how the study size was arrived at. | 8-10 |
| Missing data | 9 | D;V | Describe how missing data were handled (e.g., complete-case analysis, single imputation, multiple imputation) with details of any imputation method. | 9-10 |
| Statistical analysis methods | 10a | D | Describe how predictors were handled in the analyses. | SM-5 |
| | 10b | D | Specify type of model, all model-building procedures (including any predictor selection), and method for internal validation. | 9-10 |
| | 10c | V | For validation, describe how the predictions were calculated. | 11 |
| | 10d | D;V | Specify all measures used to assess model performance and, if relevant, to compare multiple models. | 10 |
| | 10e | V | Describe any model updating (e.g., recalibration) arising from the validation, if done. | NA |
| Risk groups | 11 | D;V | Provide details on how risk groups were created, if done. | NA |
| Development vs. validation | 12 | V | For validation, identify any differences from the development data in setting, eligibility criteria, outcome, and predictors. | 11, SM-4,18 |

| Results | | | | |
|---|---|---|---|---|
| Participants | 13a | D;V | Describe the flow of participants through the study, including the number of participants with and without the outcome and, if applicable, a summary of the follow-up time. A diagram may be helpful. | 13, SM-4 |
| | 13b | D;V | Describe the characteristics of the participants (basic demographics, clinical features, available predictors), including the number of participants with missing data for predictors and outcome. | SM-20 |
| | 13c | V | For validation, show a comparison with the development data of the distribution of important variables (demographics, predictors and outcome). | SM-28 |
| Model development | 14a | D | Specify the number of participants and outcome events in each analysis. | 13-15 |
| | 14b | D | If done, report the unadjusted association between each candidate predictor and outcome. | NA |
| Model specification | 15a | D | Present the full prediction model to allow predictions for individuals (i.e., all regression coefficients, and model intercept or baseline survival at a given time point). | 29 |
| | 15b | D | Explain how to the use the prediction model. | 2 |
| Model performance | 16 | D;V | Report performance measures (with CIs) for the prediction model. | 28-30 |
| Model-updating | 17 | V | If done, report the results from any model updating (i.e., model specification, model performance). | NA |
| **Discussion** | | | | |
| Limitations | 18 | D;V | Discuss any limitations of the study (such as nonrepresentative sample, few events per predictor, missing data). | 21 |
| Interpretation | 19a | V | For validation, discuss the results with reference to performance in the development data, and any other validation data. | 18,20 |
| | 19b | D;V | Give an overall interpretation of the results, considering objectives, limitations, results from similar studies, and other relevant evidence. | 20-21 |
| Implications | 20 | D;V | Discuss the potential clinical use of the model and implications for future research. | 21 |
| **Other information** | | | | |
| Supplementary information | 21 | D;V | Provide information about the availability of supplementary resources, such as study protocol, Web calculator, and data sets. | 2 |
| Funding | 22 | D;V | Give the source of funding and the role of the funders for the present study. | 2 |

*Items relevant only to the development of a prediction model are denoted by D, items relating solely to a validation of a prediction model are denoted by V, and items relating to both are denoted D;V. We recommend using the TRIPOD Checklist in conjunction with the TRIPOD Explanation and Elaboration document.

SM – supplementary material, NA – not applicable.
